# Reduced locus coeruleus integrity linked to response inhibition deficits in parkinsonian disorders

**DOI:** 10.1101/2021.10.14.21264996

**Authors:** Rong Ye, Frank H. Hezemans, Claire O’Callaghan, Kamen A. Tsvetanov, Catarina Rua, P. Simon Jones, Negin Holland, Maura Malpetti, Alexander G. Murley, Roger A. Barker, Caroline H. Williams-Gray, Trevor W. Robbins, Luca Passamonti, James B. Rowe

## Abstract

Parkinson’s disease and progressive supranuclear palsy (PSP) both impair response inhibition, exacerbating impulsivity. Inhibitory control deficits vary across individuals, and have been linked with worse prognosis and lack of improvement on dopaminergic therapy. Motor and cognitive control are associated with noradrenergic innervation of the cortex, arising from the locus coeruleus noradrenergic system. Here we test the hypothesis that loss of structural integrity of the locus coeruleus explains response inhibition deficits in progressive supranuclear palsy and Parkinson’s disease. This cross-sectional observational study recruited 24 people with idiopathic Parkinson’s disease, 14 with PSP-Richardson’s syndrome, and 24 age- and sex-matched controls. All participants undertook a stop-signal task and ultrahigh field 7T-magnetic transfer weighted imaging of the locus coeruleus. Hierarchical Bayesian estimation of the parameters of ‘race models’ of go-versus stop-decisions was used to quantify the cognitive processes of response inhibition. We tested the multivariate relationship between locus coeruleus integrity and model parameters using partial least squares. Both disorders impaired response inhibition at the group level. Progressive supranuclear palsy caused a distinct pattern of abnormalities in inhibitory control, relative to Parkinson’s disease and healthy controls, with a paradoxically reduced threshold for go responses, but longer non-decision times, and more lapses of attention. The variation in response inhibition correlated with variation in the integrity of the locus coeruleus, across participants in both clinical groups. Structural imaging of the locus coeruleus, coupled with behavioural modelling in parkinsonian disorders, confirms that locus coeruleus integrity is associated with response inhibition and its degeneration contributes to neurobehavioural changes. The noradrenergic system is therefore a promising target to treat impulsivity in these conditions. The optimisation of noradrenergic treatment is likely to benefit from stratification according to locus coeruleus integrity.

## Introduction

Progressive supranuclear palsy (PSP) and Parkinson’s disease (PD) have distinct neuropathology and clinical features ^1,2^, but they can both cause cognitive and behavioural problems ^3^. One of the consequences is an impairment of response inhibition, contributing to disinhibited behaviour and impulsivity. These are associated with poor clinical outcomes ^4-6^. Impulsivity is a multi-faceted behavioural construct, including abnormal sensitivity to reward, intolerance to delayed reward and a failure to inhibit inappropriate responses ^7,8^. These deficits are mediated by abnormalities in complementary frontostriatal neural circuits and neurochemical systems ^9^. Here we focus on response inhibition, as the execution of responses represents a point of convergence for upstream changes in cognition and behavioural decisions. Deficits in inhibitory control arise from pathology in frontostriatal circuits, including the ventrolateral prefrontal cortex, pre-motor cortex, caudate and sub-thalamic nuclei, and their monoaminergic regulation. Noradrenergic manipulations influence response inhibition, especially the cancellation of a response, in preclinical models^10-12^, healthy humans^13^, attention deficit disorders^14,15^ and Parkinson’s disease^16-20^. They are therefore a promising route to ameliorate response inhibition deficits in diverse neurological and psychiatric disorders.

The locus coeruleus is the main source of noradrenaline in the brain and an early site of pathology in progressive supranuclear palsy and Parkinson’s disease ^21-24^. Locus coeruleus degeneration is severe in symptomatic progressive supranuclear palsy and Parkinson’s disease, although *post mortem* studies indicate a high degree of variability in locus coeruleus cell loss ^24,25^. The heterogeneity in locus coeruleus damage has been linked to variability in the response to drugs that increase noradrenergic transmission such as atomoxetine ^26^. To facilitate more targeted treatment of impulsivity in progressive supranuclear palsy and Parkinson’s disease, it is necessary to quantify locus coeruleus structural integrity *in vivo* and determine its relationship to response inhibition. Specialist magnetic resonance imaging (MRI) sequences for ultrahigh field scanners (7T) have enabled sensitive and well-tolerated quantification of locus coeruleus pathology ^25-28^. The resolution is sufficient to examine regional effects of pathology within the locus coeruleus ^29,30^.

Noradrenergic deficits may influence response inhibition at both motor and cognitive (decisional) levels, given their widespread cortical projections ^31^. Response inhibition can be measured using performance on a stop signal task, but singular parameters of performance may obscure the complexity of underlying decision mechanisms ^9,16,18^. A multivariate model of computational parameters of response inhibition overcomes this limitation, to distinguish motor, attentional and decisional components of inhibition ^26,32,33^.

In this study, we used complementary parametric analyses of the stop signal task, estimated with hierarchical Bayesian models, to advance our understanding of the response inhibition deficits in progressive supranuclear palsy and Parkinson’s disease. The first model explains response accuracy and reaction times as function of a race between three processes: a stop process, a go process for the response that matches the choice stimulus, and a go process for the response that mismatches the choice stimulus ^34-36^. To decompose the cognitive mechanisms of response inhibition, we also parameterised accumulation-to-threshold models in the second type of model, in which evidence accumulates stochastically until a go response threshold is reached. The mean time required to trigger the stop process served as the estimate of the stop signal reaction time (SSRT). The models also estimate attentional failures to trigger the stop and go processes.

We tested the hypothesis that these parameters of response inhibition relate to locus coeruleus structural integrity at an individual patient level, as measured *in vivo* using 7T MRI. We predicted that the response inhibition deficits characterising progressive supranuclear palsy and Parkinson’s disease would be associated with reduced locus coeruleus integrity. Given the multivariate nature of the model parameters and the topographic organisation of the locus coeruleus, we investigated the multivariate relationship between locus coeruleus integrity and response inhibition.

## Methods and Materials

### Participants

Fourteen patients with probable PSP-Richardson’s syndrome (MDS 2017 criteria), 24 with idiopathic Parkinson’s disease (UK Parkinson’s disease Brain Bank criteria), and 24 age- and sex-matched healthy controls were included in the study. Controls did not use psychoactive medications and exclusions criteria for all participants included history of stroke, severe medical co-morbidity, and any contraindications to 7T MRI. None of the patients met criteria for impulse control disorders, based on clinical impression and/or the Questionnaire for Impulsive-Compulsive Disorders. Participants were not demented, based on a Mini-Mental State Examination score >26 and clinical impression. All patients with Parkinson’s disease and 10/14 progressive supranuclear palsy patients were on dopaminergic medications (Table 1). Eighteen patients with Parkinson’s disease were part of a single-dose, placebo-controlled, crossover drug study where the relationship between locus coeruleus integrity and drug responsiveness was investigated ^26^. The behavioural performance of these patients was examined for the placebo session, and potential placebo or practice effects were explicitly modelled (see **Statistical Analyses**). All participants underwent a structured clinical, cognitive, and behavioural assessment (Table 1). The study was approved by the local Cambridge Research Ethics Committees. Participants provided written informed consent according to the Declaration of Helsinki.

**Table 1.** Demographics (mean and standard deviation) of participants and clinical assessments.

|  | Descriptive |  |  | P values for pairwise tests |  |  |
| --- | --- | --- | --- | --- | --- | --- |
|  | HC | PD | PSP | HC vs PD | HC vs PSP | PD vs PSP |
| Age (years) | 65.5 (5.5) | 67.2 (7.4) | 69.7 (7.7) | 0.66 | 0.164 | 0.52 |
| Education (years) | 14.8 (3.1) | 14 (2.3) | 12.3 (2.8) | 0.584 | 0.021 | 0.15 |
| Male/Female | 13/11 | 18/6 | 8/6 | 0.131 | 0.859 | 0.253 |
| MMSE | 29.75 (0.53) | 29.52 (0.65) | 28.5 (1.74) | 0.649 | **<0.001** | 0.009 |
| MoCA | 28.58 (1.44) | 27.88 (1.87) | 24 (3.94) | 0.557 | **<0.001** | **<0.001** |
| ACER-total | 97.71 (3.25) | 95.25 (3.6) | 87.21 (7.17) | 0.153 | **<0.001** | **<0.001** |
| Apathy Scale | 10.38 (5.25) | 12.42 (5.55) | 20 (9.49) | 0.528 | **<0.001** | 0.003 |
| BIS | 55.71 (9.56) | 58.69 (10.21) | 63.86 (12.44) | 0.591 | 0.063 | 0.316 |
| HADS-depression | 2.83 (2.84) | 4.25 (2.79) | 7.43 (4.27) | 0.281 | **<0.001** | 0.012 |
| HADS-anxiety | 4.29 (3.53) | 5.17 (3.16) | 6.57 (3.2) | 0.634 | 0.111 | 0.424 |
| RBDSQ | - | 5.38 (3.69) | 3.07 (1.82) | - | - | 0.036 |
| Disease Duration (years) | - | 5.09 (3.05) | 4.24 (2.68) | - | - | 0.397 |
| LEDD | - | 646.6 (509.53) | 323.57 (389.4) |  |  | 0.038 |
| UPDRS-III | - | 28.21 (12.21) | 33.07 (6.96) | - | - | 0.182 |
| PSPRS | - | - | 30.79 (9.11) | - | - | - |

Group difference in sex was examined using chi-square test. A one-way ANOVA was used for group difference with post-hoc Tukey HSD p values provided for pairwise comparisons. RBDSQ, disease duration and UPDRS-III were compared with independent samples t-test between PD and PSP. MMSE: Mini-Mental State Examination, MoCA: Montreal Cognition Assessment, ACER: Addenbrooke's Cognitive Examination Revised, BIS: Barratt Impulsiveness Scale, HADS: Hamilton Anxiety and Depression Scale, RBDSQ: REM Sleep Behaviours Screening Questionnaire, LEDD: Levodopa equivalent daily dose, UPDRS: Unified Parkinson's Disease Rating Scale, PSPRS: Progressive Supranuclear Palsy Rating Scale. Significant P values ( $P < 0.0016$ , equivalent to $P < 0.05$ with Bonferroni correction) are highlighted in bold.

### Stop-signal task

Response inhibition was measured using a stop-signal paradigm where a two-choice reaction time (RT) ‘go’ task was occasionally interrupted by a ‘stop signal’. For the go task, participants were instructed to indicate the direction of a black arrow presented at the centre of the screen by pressing a left or right button. On stop trials, the arrow changed to a red colour in conjunction with a tone (i.e., the stop signal) and participants were instructed to withhold their initiated response (**Fig 1, top-left**). The stop signal occurred after a variable delay (i.e., the stop-signal delay, SSD), the length of which was determined by an adaptive staircase method. The SSD ranged from 50 ms to 1500 ms and increased or decreased by 50 ms after a successful or failed stop trial, respectively. Further details of the task design are provided in O’Callaghan et al. ^26^.

**Figure 1.**
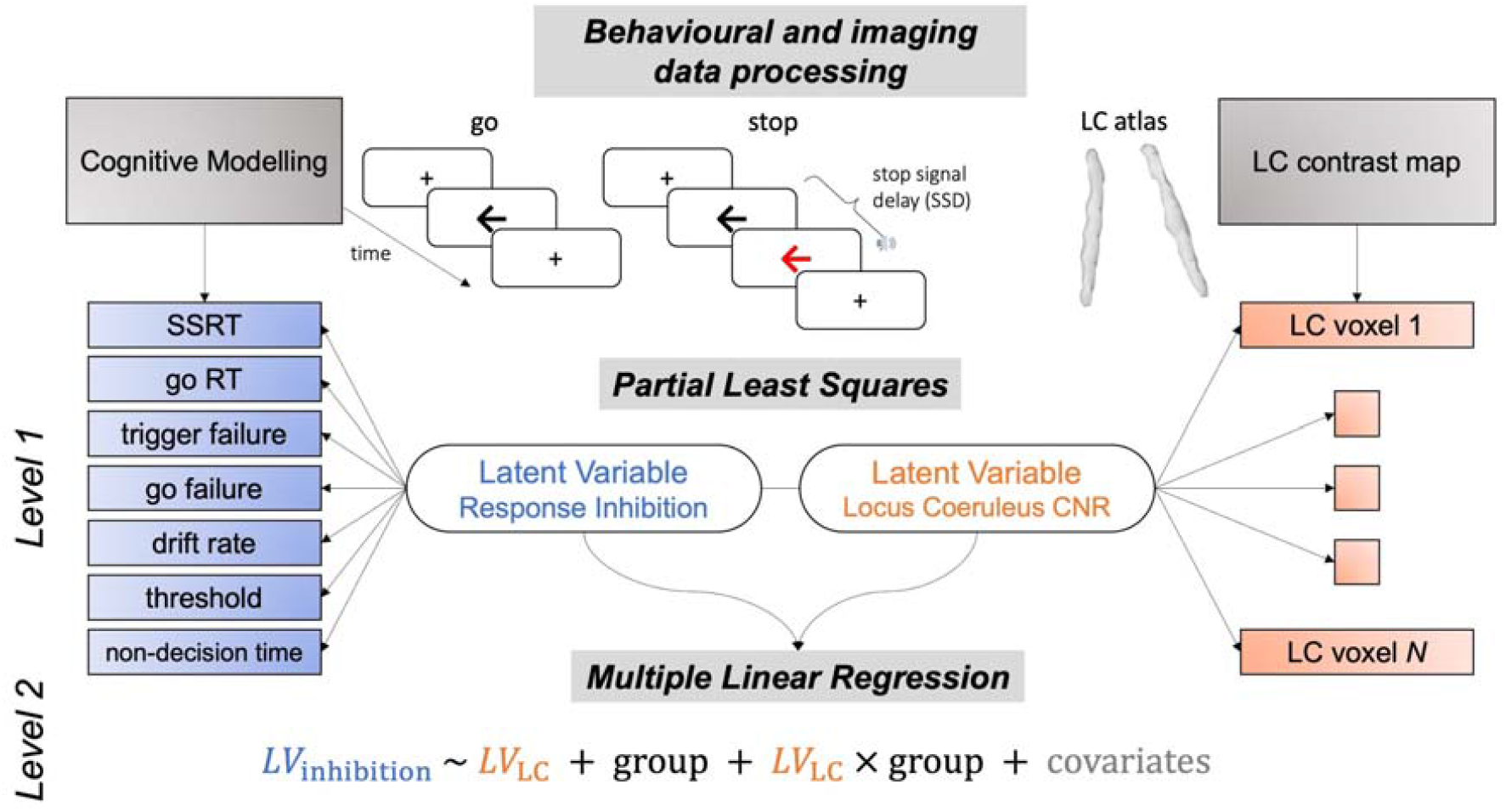
Schematic representation of data analysis pipeline. The trial-by-trial stop signal task performance was subjected to a two parametric race model following ex-Gaussian and shifted Wald distributions. An array of behavioural parameters were estimated hierarchically from the models for both stop and go response, including stop-signal reaction time (SSRT), go reaction time (go RT), trigger failure, go failure, drift rate (v), response threshold (B) and non-decision time (t0). These parameters altogether provided more mechanistic understanding of response inhibition. The locus coeruleus (LC) integrity was assessed by computing voxel-wise contrast-to-noise ratio (CNR) and extracted using an independent LC probability atlas. The multivariate relationship between LC integrity and response inhibition was then examined using partial least squares using resulted behavioural and imaging matrices from previous data processing steps. Significant pairs of latent variables were identified with the permutation test. The contribution of LC in response inhibition was finally confirmed in linear regression models with individual subject loading scores on the inhibition latent variable as dependent variable, loading scores on the LC latent variable, group and nuisance covariates as predictors.

### Modelling of response inhibition

#### Parametric race models

We applied two complementary models to the stop-signal task data. The first model assumed that the finish time distributions of the stop and go processes followed ex-Gaussian distributions ^37^, with mean µ and standard deviation σ of the Gaussian component, and mean τ of the exponential component. The mean finish time of each process was estimated as the mean of the corresponding ex-Gaussian distribution, which is given by µ + τ. The mean finish times of the stop process and matching go process were taken as the stop signal reaction time (SSRT) and go RT, respectively.

The second model was similar, except that the finish time distributions of the go processes were assumed to follow shifted Wald distributions ^36^. The Wald distribution describes the first-passage time distribution of a single-boundary diffusion process, where evidence accumulates stochastically at a positive mean rate (cf. the drift rate) until a threshold is reached. A subject-specific constant non-decision time shifts the lower bound of the distribution to account for peripheral processes, such as stimulus encoding and motor output. The shifted Wald distribution therefore enables a process model that explains how the go RT distributions were generated.

Both models additionally included the probabilities of attentional failures related to the stop process (trigger failure) and go processes (go failure). Thus, the first model featured 11 free parameters: three ex-Gaussian parameters for each of the three processes, and the trigger and go failure probabilities. The second model featured 9 free parameters: separate drift rates for the matching and mismatching go processes, a threshold and non-decision time that were shared across the go processes, three ex-Gaussian parameters for the stop process, and the trigger and go failure probabilities.

Hierarchical Bayesian modelling was used to fit each model to the observed task data, separately for the progressive supranuclear palsy, Parkinson’s disease, and control groups. Markov Chain Monte Carlo (MCMC) sampling methods were used to estimate the posterior distributions of all group- and participant-level parameters. We assigned broad (“weakly informative”) priors on the group-level means and standard deviations of the model parameters (**Table S1**). Sampling convergence was confirmed by visual inspection of the time series plots of the MCMC samples, and by the potential scale reduction statistic R-hat (< 1.1 for all parameters). The absolute goodness of fit was assessed by visually comparing the observed data to simulated data generated from the model’s posterior predictive distribution (**Fig.S5-7**).

Model fitting was performed using the Dynamic Models of Choice (DMC) toolbox ^38^ implemented in R (version 3.6.1). The number of sampling chains was set to three times the number of free parameters. Automated procedures were used to continue sampling until convergence was reached (h.run.unstuck.dmc and h.run.converge.dmc functions in the DMC toolbox). After this, an additional 500 iterations were obtained for each chain to create a final posterior distribution for each parameter, which was used for statistical analyses.

### MRI acquisition and processing

Participants were scanned on a 7T Magnetom Terra (Siemens, Erlangen, Germany) with a 32-channel receive head coil (Nova Medical, Wilmington, USA). Following the acquisition and processing protocol described previously ^26,27^, the locus coeruleus was imaged using a near-isotropic 3-D magnetisation transfer (MT) weighted sequence at submillimetre resolution (0.4 × 0.4 × 0.5 mm^3^, 112 oblique, axial slices oriented perpendicular to the long axis of the brainstem). Two images with MT presaturation pulses were acquired then averaged off-line to enhance signal-to-noise ratio. One image without MT effect (MT-off) was additionally obtained for registration. A high resolution T1-weighted structural image (0.7 mm isotropic) was acquired using an MP2RAGE sequence with the UK7T Network harmonised protocol ^39^: TE = 2.58 ms, TR = 3500 ms, BW = 300 Hz/px, voxel size = 0.7 × 0.7 × 0.7 mm^3^, FoV = 224 × 224 × 157 mm^3^, acceleration factor (A>>P) = 3, flip angles = 5/2°. MT images were bias field corrected then co-registered to the isotropic 0.5 mm ICBM152 (International Consortium for Brain Mapping) T1-weighted asymmetric nonlinear template ^40^ following a T1-driven coregistration approach using the Advanced Normalization Tools (ANTs v2.2.0). The individual registration roadmap was initiated from the estimation between averaged MT image and the MT-off image then moved to the coregistration between MT and T1 modality in the following order: MT-off to MT, individual T1 to MT-off, individual T1 to T1 group template and finally T1 group template to the ICBM152 template.

The co-registered MT images were converted to contrast-to-noise (CNR) maps by subtracting the mean and dividing by the standard deviation of the signal in a central pontine reference region ^26,27^. A probabilistic locus coeruleus atlas was applied on the CNR maps with a conservative threshold (25%) to extract voxel-wise locus coeruleus CNRs for later statistical tests and slice-wise means for group comparisons. Structural T1-weighted images were subjected to FreeSurfer (v 6.0) *recon-all* pipeline with *-highres* and *-brainstem* options. The resulting total intracranial and brainstem volumes were used for the estimation of global and local atrophy, respectively.

### Statistical Analysis

#### Group differences in response inhibition parameters

The go error rate was defined as the proportion of go trials with an incorrect response, including commission errors (responses that mismatched the choice stimulus) and omission errors (missing responses). The stop accuracy rate was defined as the proportion of stop trials with successfully inhibited (i.e. missing) responses. Each of these measures served as dependent variables in an ANOVA with group as the between-subjects factor. Differences between specific groups were then examined with post-hoc Tukey’s tests.

We focused on group differences by examining the posterior distributions of the group-level means of the race model parameters. For each posterior distribution, we took the median as the posterior estimate, and the 95% quantile interval (QI) as the range of plausible values. We derived posterior distributions for group contrasts by subtracting the set of MCMC samples of the two groups under consideration. For each group contrast, we computed the probability of direction (*P*_dir_) as an index of the presence of an effect ^41^. This measure indicates the proportion of the contrast’s posterior distribution that is strictly positive or negative (whichever is the most probable). Note that *P*_dir_ can be directly interpreted as the probability that a group difference is non-zero, and is therefore not subject to a particular significance threshold. To examine individual differences in mechanisms of response inhibition, we extracted the medians of all participant-level posterior distributions.

#### The relationship between locus coeruleus integrity and response inhibition

We studied the relationship between locus coeruleus integrity and response inhibition deficits with a two-level analytical strategy ^42,43^. First, the multidimensional relationship was examined using partial least squares (PLS) ^42,44^ with in-house Matlab (R2018b) scripts where pairs of latent variables were computed from all response inhibition parameters (LV_inhibition_) and voxelwise locus coeruleus contrast (LV_LC_). The PLS multivariate method is particularly suitable given the ratio of variables to participants ^45^. The significant pair of latent variables was identified using a permutation test (10,000 iterations, *P* < 0.05).

Second, individual loadings on LVs were subjected to linear regression models to confirm the group-wise relationship between locus coeruleus integrity and response inhibition ability. Subsequent regression models examined the effects of nuisance covariates including age, global and local atrophy, disease duration and motor severity. To mitigate the potential placebo and/or practice effects in the 18/24 of the patients with Parkinson’s disease who had completed the task as part of a drug study ^26^, we adopted two approaches using (i) a categorical variable indicating session (0 = first / only session; 1 = second session) and (ii) a categorical indicator for drug order (0 = one session without placebo; 1 = first session on placebo; 2 = second session on placebo). Adding these categorical indicators as a covariate of no interest in the regression models did not meaningfully change the relationship between locus coeruleus integrity and response inhibition (see results). Prior to analysis, continuous variables were z-scored and categorical variables (including nuisance covariates) were assigned sum-to-zero contrasts.

## Results

### Basic task performance

Participant characteristics and clinical summary data are presented in **Table 1**. Groups were similar by age, sex and education, while the patient groups were similar by disease duration and motor severity, although cognitive function was lower in progressive supranuclear palsy. There were expected significant main effects of group on both the stop accuracy rate (**Fig.2A**; *F*_(2, 59)_ = 10.21, *P* < .001; *BF* = 143.66) and go error rate (**Fig.2D**; *F*_(2, 59)_ = 8.46, *P* < .001; *BF* = 46.85). Post hoc tests indicated that these effects were driven by patients with progressive supranuclear palsy, who had reduced response accuracy compared to the other groups.

**Figure 2.**
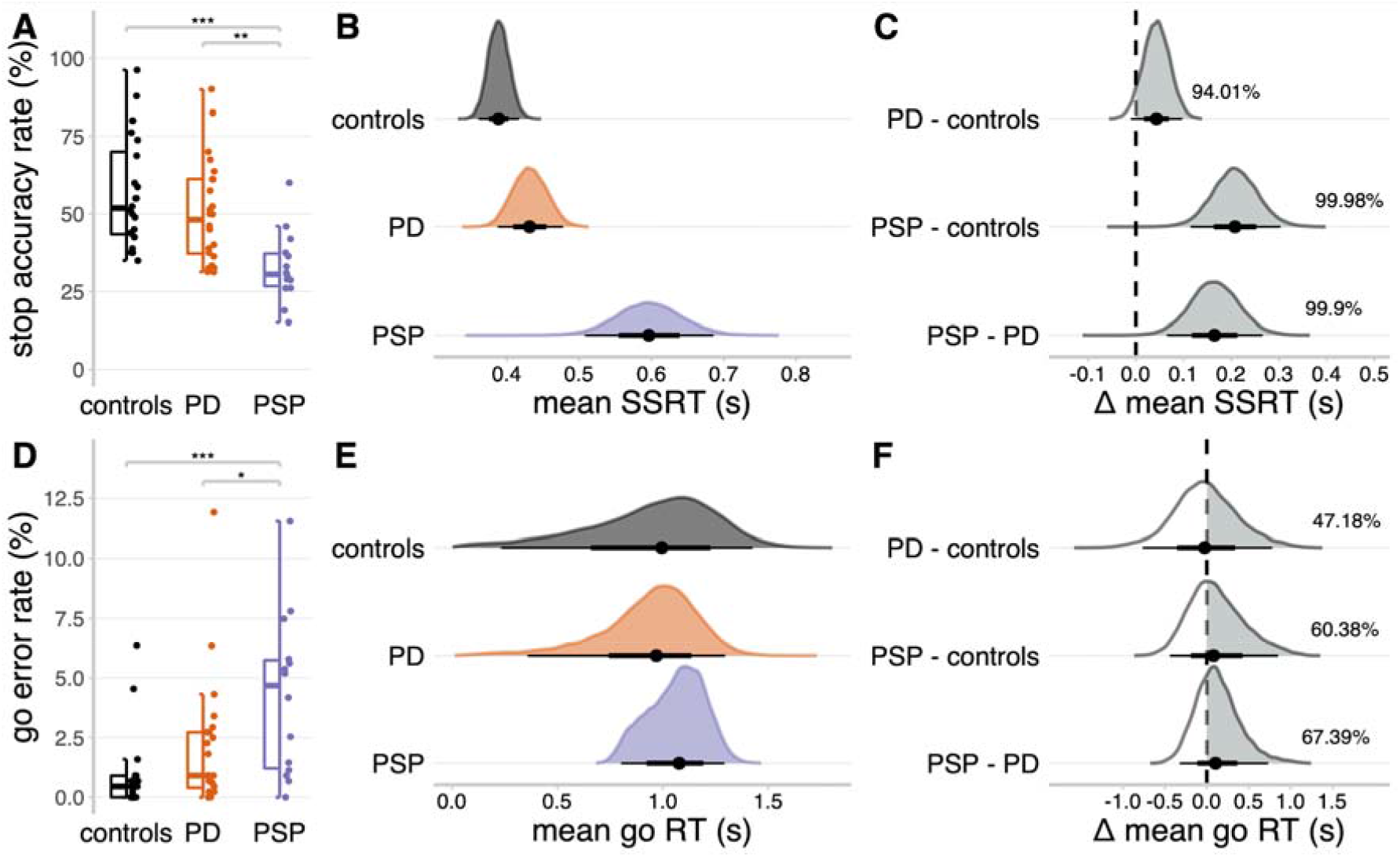
Stop signal task performance. (A, D): Proportions of successful stop trials (A) and incorrect go responses (D). Asterisks represent the statistical significance of post-hoc Tukey’s tests comparing groups: ***, *P* < .001; **, *P* < .01; *, *P* < .05. (B, E): Posterior distributions of the mean stop signal reaction times (B) and mean go reaction times (E). The black dots represent the medians; thick black line segments represent the 66% quantile intervals; thin black line segments represent the 95% quantile intervals. (C, F): Posterior distributions of group comparisons for the mean stop signal reaction time (C) and mean go reaction time (F). Percentages indicate the proportion of the posterior distribution that is strictly positive, i.e. the grey shaded area.

The mean go RT (**Fig.2E**) was similar across progressive supranuclear palsy (median = 1.08 s, 95% QI: [0.80, 1.29]), Parkinson’s disease (median = 0.97 s, 95% QI: [0.36, 1.30]), and control (median = 1.00 s, 95% QI: [0.23, 1.43]) groups (**Fig.2F**). However, the mean SSRT was longer in progressive supranuclear palsy (**Fig.2B**; median = 0.60 s, 95% QI: [0.51, 0.69]) compared to controls (**Fig.2C**; median = 0.39 s, 95% QI: [0.36, 0.42]; Δ PSP - controls: median = 0.21 s, 95% QI: [0.11, 0.30], *P*_dir_ = 99.98%) and Parkinson’s disease (median = 0.43 s, 95% QI: [0.39, 0.48]; Δ PSP - PD: median = 0.16 s, 95% QI: [0.06, 0.27], *P*_dir_ = 99.90%), confirming the expected impairment of response inhibition. The SSRT was also longer in Parkinson’s disease compared to controls (**Fig.2B**; Δ PD – controls: median = 0.04 s, 95% QI: [−0.01, 0.10], *P*_dir_ = 94.01%).

### Processes underlying response inhibition deficits

Following Zhang et al (2015), the mean threshold height was confirmed as lower in progressive supranuclear palsy (**Fig.3A**; median = 2.17, 95% QI: [1.93, 2.40]) relative to controls (**Fig.3B**; median = 3.31, 95% QI: [2.69, 3.80]; Δ PSP - controls: median = −1.14, 95% QI: [−1.68, −0.49], *P*_dir_ = 99.71%) and Parkinson’s disease (median = 3.02, 95% QI: [2.38, 3.53]; Δ PSP - PD: median = −0.85, 95% QI: [−1.41, −0.18], *P*_dir_ = 98.99%). The non-decision time was slower in progressive supranuclear palsy (**Fig.3C**; median = 0.22 s, 95% QI: [0.11, 0.34]) relative to controls (**Fig.3D**; median = 0.11 s, 95% QI: [0.10, 0.13]; Δ PSP - controls: median = 0.11 s, 95% QI: [−0.004, 0.23], *P*_dir_ = 96.37%) and Parkinson’s disease (median = 0.12 s, 95% QI: [0.10, 0.16]; Δ PSP - PD: median = 0.10 s, 95% QI: [−0.02, 0.22], *P*_dir_ = 93.67%). However, the posterior estimate of progressive supranuclear palsy patients’ non-decision time was imprecise (**Fig.3C**), suggesting that a slower non-decision time was not a feature across all patients with progressive supranuclear palsy.

**Figure 3.**
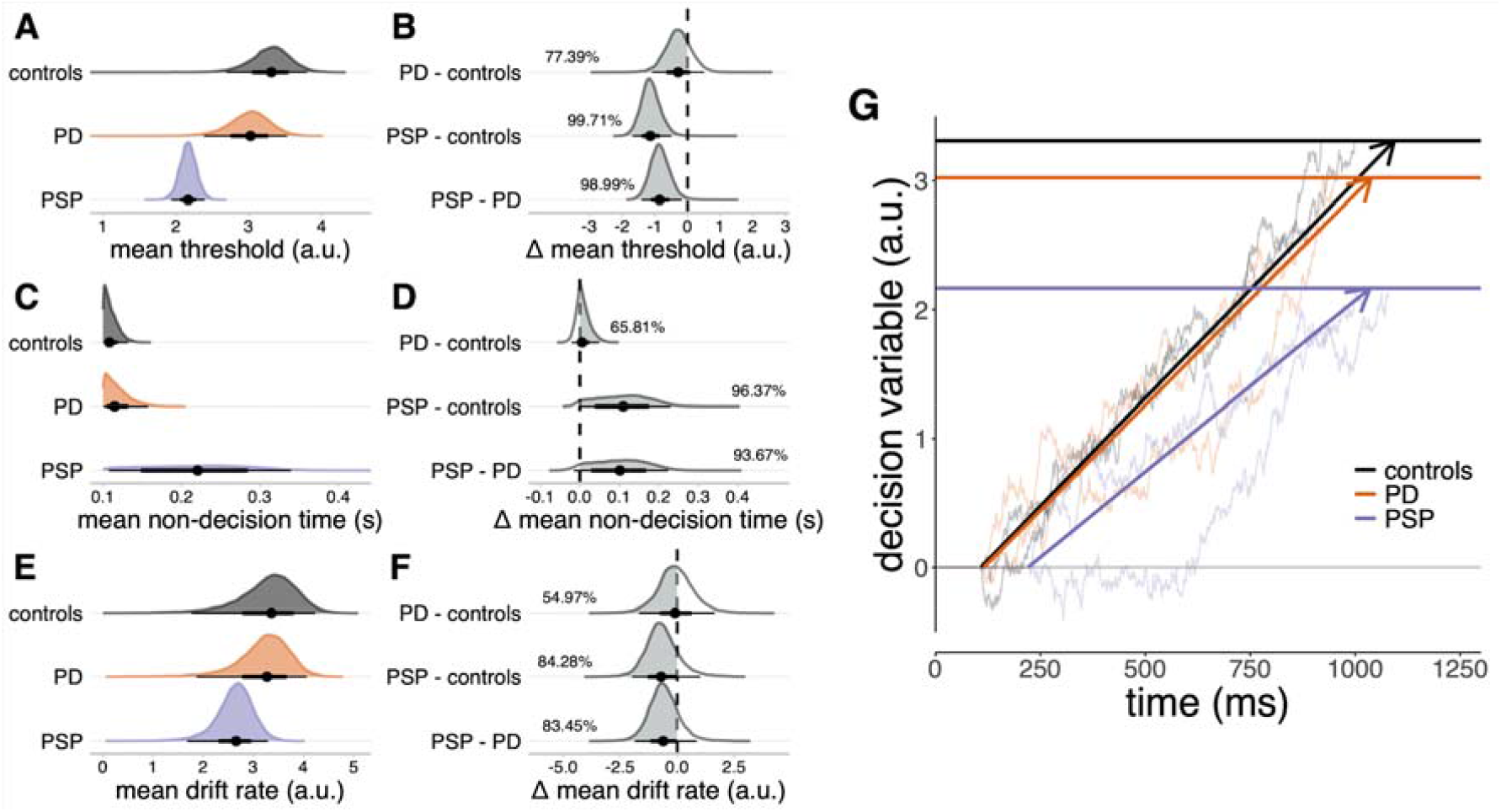
Sequential sampling model of go responses. (A, C, E): Posterior distributions of mean threshold (A), mean non-decision time (C), and mean drift rate (E). The black dots represent the medians; thick black line segments represent the 66% quantile intervals; thin black line segments represent the 95% quantile intervals. (B, D, E): Posterior distributions of group comparisons for the mean threshold (B), mean non-decision time (D), and mean drift rate (F). Percentages indicate the proportion of the posterior that is strictly negative (B, F) or positive (D), i.e. the grey shaded area. (G) Schematic illustration of the sequential sampling model. Evidence for a go response accumulates stochastically at a constant average rate (the drift rate) until a threshold is reached. To explain reaction times, the time taken by the accumulator to reach the threshold is offset by a constant non-decision time. The bold horizontal lines represent the posterior median estimates of the mean thresholds. The bold directed lines represent the posterior median estimates of the mean drift rate. The thin lines provide examples of simulated random walks.

We estimated the probabilities of failing to trigger the stop process (trigger failure) and go processes (go failure). The mean probit-transformed trigger failure probability was higher in progressive supranuclear palsy (**Fig.4A**; median = −1.78, 95% QI: [−2.68, −0.87]) compared to controls (**Fig.4B**; median = −3.06, 95% QI: [−3.71, −2.40]; Δ PSP - controls: median = 1.27, 95% QI: [0.18, 2.40], *P*_dir_ = 98.85%). The mean probit-transformed go failure probability was higher in the progressive supranuclear palsy group **(Fig.4C;** median = −2.78, 95% QI: [−3.12, −2.40]) compared to the control (**Fig.4D**; median = −3.74, 95% QI: [−4.13, −3.35]; Δ PSP - controls: median = 0.96, 95% QI: [0.45, 1.49], *P*_dir_ = 99.98%) and Parkinson’s disease group (median = −3.46, 95% QI: [−3.81, −3.12]; Δ PSP - PD: median = 0.69, 95% QI: [0.21, 1.19], *P*_dir_ = 99.76%). There were no meaningful differences between the Parkinson’s disease and control groups on these attentional parameters (**Fig.4B&D**; all *P*_dir_ < 90%).

**Figure 4.**
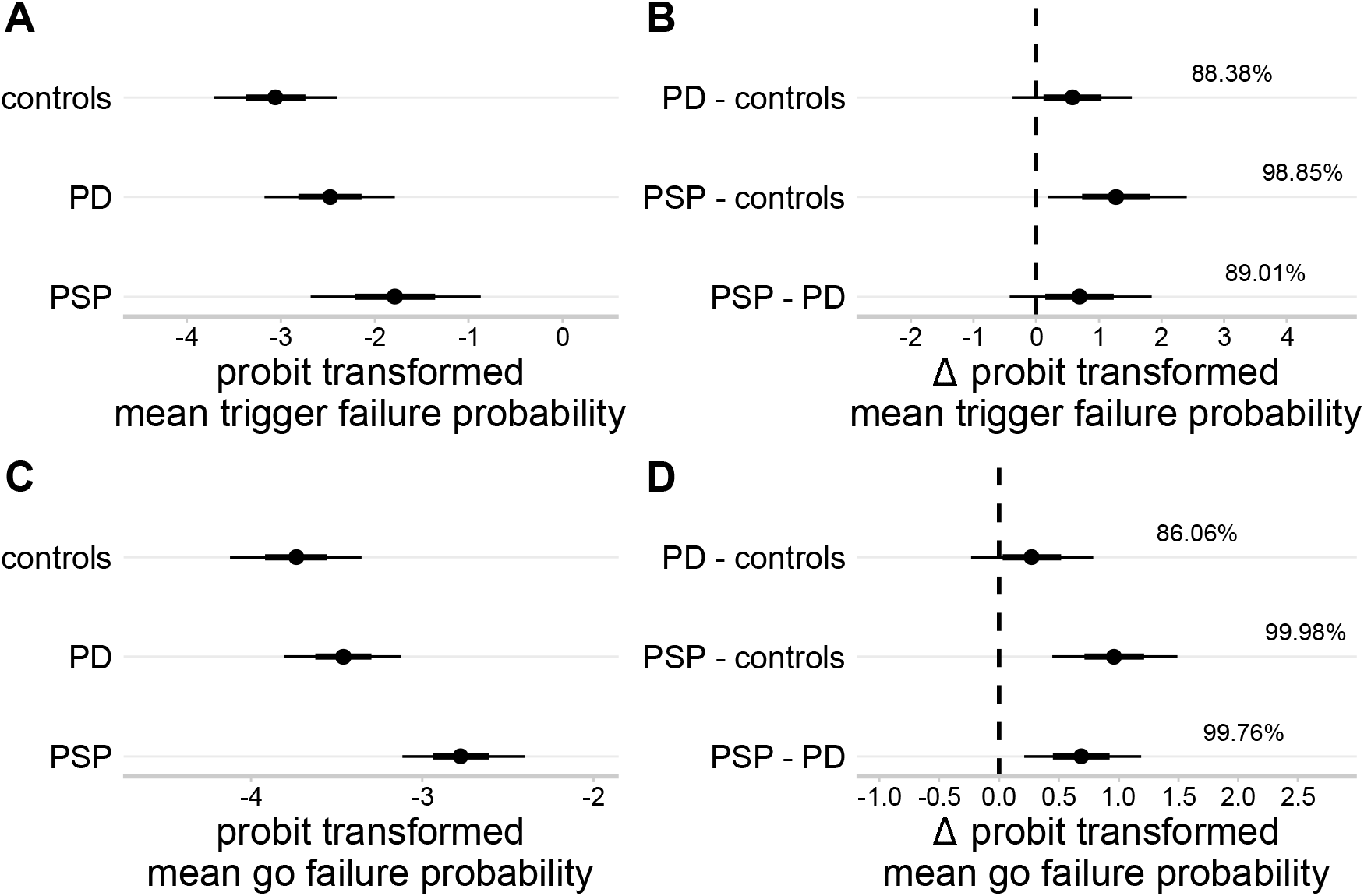
Group-level means of attentional failure parameters. (A, C): Posterior distributions of probit transformed mean trigger failure probability (A), and probit transformed mean go failure probability (C). The black dots represent the medians; thick black line segments represent the 66% quantile intervals; thin black line segments represent the 95% quantile intervals. (B, D): Posterior distributions of group comparisons for the probit transformed mean trigger failure probability (B) and probit transformed mean go failure probability (D). Percentages indicate the proportion of the posterior that is strictly positive, i.e. the grey shaded area. Note that the probit function was used to project the attentional failure parameters from the probability scale (0, 1) to the real line (-∞, ∞).

### Locus coeruleus integrity and individual differences

The locus coeruleus integrity was reduced in the caudal region for both patient groups (**Fig.S1-S4**; **Fig.5C**). Partial least squares analysis (PLS) of the response inhibition parameters and voxel-wise locus coeruleus contrast identified a single significant pair of latent variables (*r* = 0.75, *P* = .040, 10,000 permutations).

**Figure 5.**
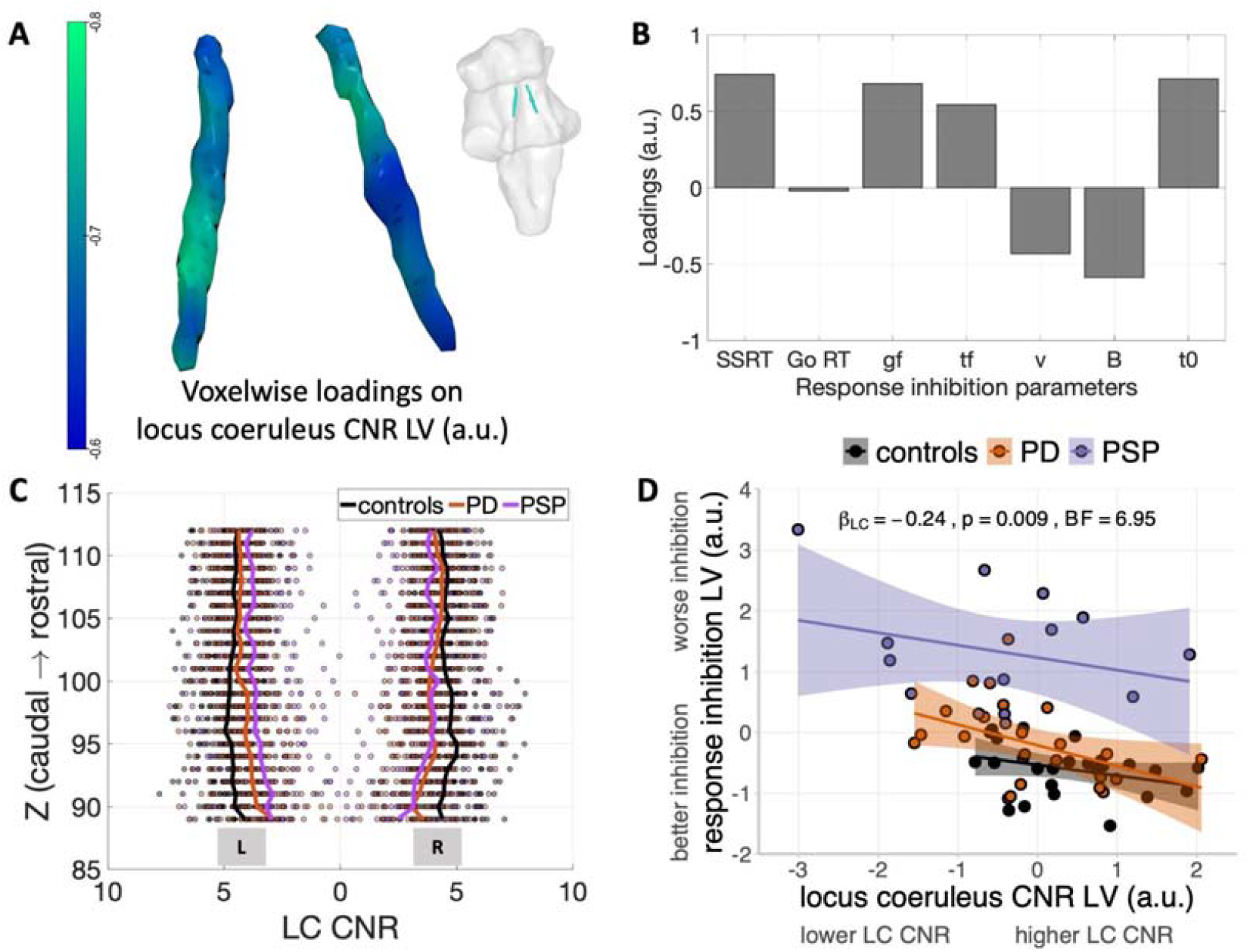
The relationship of locus coeruleus integrity and response inhibition. As confirmed using partial least squares, a significant pair of latent variables was identified between voxel-wise LC contrast and response inhibition parameters estimated from cognitive models. The spatial distribution of voxel-wise LC loadings (A) and individual loadings on response inhibition latent variable (B) were presented. Negative LC loadings were associated with positive loadings on stop-signal reaction time (SSRT), go failure (gf), trigger failure (tf) and non-decision time (t0), and negative loadings on drift rate (v) and response threshold (B). This suggested that impaired response inhibition is linked with reduced LC integrity seen in both PD and PSP patients (C). An overall relationship of LC integrity and response inhibition was further confirmed in the linear regression model consistent across all groups as supported by a significant main effect of LC when including the group predictor in the model (D). Individual fitted lines for each group were presented with curved areas indicating 95% confidence intervals.

The locus coeruleus latent variable (LV_LC_) expressed negative loadings throughout the structure, to a variable degree across sub-regions (**Fig.5A**). The response inhibition latent variable (LV_inhibition_) expressed positive loadings on the SSRT, go and trigger failure probabilities, and non-decision time, and expressed negative loadings on the drift rate and response threshold of the go process (**Fig.5B**). Thus, higher scores on this behavioural latent variable reflected impaired response inhibition with prolonged SSRT, greater probability of attentional failures, reduced response threshold, lower drift rate, and longer non-decision time. Mean go RTs had negligible loading on the LV_inhibition_, confirming that go RTs were not related to the cognitive mechanisms of response inhibition.

A regression analysis examined the relationship between response inhibition and locus coeruleus integrity, and potential group differences. The LV_inhibition_ participant score was the dependent variable, and the LV_LC_ participant score, group, and their interaction were independent variables. There was a significant overall relationship between the LV_LC_ scores and the LV_inhibition_ scores (**Fig.5D;** ß = −0.24, F(1, 56) = 7.23, *P* = .009; BF = 6.95). This suggests that individuals with reduced locus coeruleus integrity have more severe deficits in response inhibition. There was a significant main effect of group on the LV_inhibition_ scores (*F*_(2, 56)_ = 29.88, *P* < .001; *BF* = 1.98 × 10^7^), reflecting impaired response inhibition in the progressive supranuclear palsy group compared to the control group (*t*_(56)_ = 7.46, *P* < .001) and PD group (*t*_(56)_ = 6.33, *P* < .001), regardless of locus coeruleus CNR. There was no significant interaction effect between the LV_LC_ scores and group (*F*_(2, 56)_ = 0.31, *P* = .734; *BF* = 0.15), suggesting that the slope between the LV_LC_ and LV_inhibition_ scores is similar across groups.

We confirmed the robustness of these results with four additional regression analyses. First, adding age, gender, and years of education to the regression model as covariates of no interest did not meaningfully change the relationship between the LV_LC_ and LV_inhibition_ scores (ß = −0.23, *F*_(1, 53)_ = 6.31, *P* = .015; *BF* = 6.99), nor did the inclusion of additional covariates, including disease duration, motor severity, and local and global brain atrophy. Model selection procedures consistently identified a relatively sparse model as the optimal account of the data, retaining only the LV_LC_ scores and group as predictors of LV_inhibition_ (**Table S2-5**). Third, re-running the regression analysis with only the Parkinson’s disease and progressive supranuclear palsy groups yielded a similar relationship between the LV_LC_ and LV_inhibition_ scores (ß = −0.26, *F*_(1, 34)_ = 4.90, *P* = .034; *BF* = 2.07; **Table S6**). Fourth, accounting for potential placebo and / or practice effects in the Parkinson’s disease group did not meaningfully change the results (**Table S7-8**).

## Discussion

This study confirms the hypothesis that response inhibition deficits in Parkinson’s disease and progressive supranuclear palsy are linked to reduced structural integrity of the locus coeruleus, the principal source of cerebral noradrenaline. Diminished response inhibition (i.e., multivariate disinhibition-related parameters) was associated with reduced locus coeruleus integrity (i.e., multivariate voxelwise loadings) across all groups, in keeping with psychopharmacological and pre-clinical studies ^10,11,16-20,46^.

The race models identified latent variables to explain the behavioural performance. Despite similar mean reaction times in the go responses, there were disease-specific patterns underlying abnormal inhibitory control. Patients with progressive supranuclear palsy had a reduced response threshold, consistent with a paradoxical bias towards committing go responses as proposed by Zhang et al. ^32^, noting that the current study was manual not oculomotor. The progressive supranuclear palsy group also had a slower non-decision time, suggesting they required more time for sensory encoding and execution of motor outputs. There was evidence for a lower drift rate among patients with progressive supranuclear palsy relative to patients with Parkinson’s disease and controls, confirming the slower accumulation of evidence to reach a decision.

Task performance might also have been influenced by attentional problems, which are a common cognitive feature in parkinsonian disorders, especially in progressive supranuclear palsy^3^. To account for this possibility, we included attentional failures related to the stop and go processes (i.e., trigger failure and go failure) in the race models of the stop signal task. These indices revealed that patients with progressive supranuclear palsy had greater attentional deficits than Parkinson’s disease and control groups, albeit not to the extent that prevented them from correctly executing the task. The attentional impairments during the Stop/Go paradigm were also captured within the main PLS-derived LV_inhibition_ variable that related to the LV_LC_ component reflecting locus coeruleus structural integrity.

The results support the hypothesis that the decision threshold is reduced in progressive supranuclear palsy to compensate for the impairment in evidence accumulation (reduced drift rate) and execution (slower non-decision time). This threshold compensation is liable to change the quality of decisions, and thereby promote impulsivity. This relationship is disease agnostic and relates to the level of pathology with the locus coeruleus which is more severely affected in progressive supranuclear palsy^25^.

The variability of locus coeruleus degeneration across patients in our study is also evident *post mortem* ^21,22,24^, consistent with what is observed in other neurodegenerative disorders such as Alzheimer’s disease, corticobasal degeneration and Lewy Body dementia ^47-50^. This highlights the potential of locus coeruleus imaging as a trans-diagnostic marker, to understand individual differences in cognition beyond the classic nosological borders and diagnostic criteria ^51^.

On average, locus coeruleus degeneration is more severe in progressive supranuclear palsy than Parkinson’s disease, and our *in vivo* imaging data confirms this, particularly in the caudal portions ^25^. This distribution is consistent with neuropathological studies reporting greater degeneration in the caudal locus coeruleus ^52^. However, the response inhibition deficits spanning progressive supranuclear palsy and Parkinson’s disease topographically map to the mid-caudal and rostral locus coeruleus, that innervate the forebrain regions associated with response inhibition and impulsive behaviour ^30^.

The locus coeruleus-noradrenergic system’s influence on response inhibition and impulsivity may be non-linear (e.g. a U-inverted shape function), and involve multiple brain networks ^10,11,53^. The locus coeruleus diffusely projects to many sites within the brain where noradrenaline has a state-dependant effect on the neuronal input-output gain function ^54^. Although we did not directly measure noradrenaline transmission or noradrenergic receptor density, locus coeruleus structural integrity is a proxy index of noradrenergic function ^51,55^.

Our study focused on noradrenergic contributions to response inhibition deficits in parkinsonian disorders. However, we recognise that noradrenergic projections from the locus coeruleus have secondary pharmacological interactions with other neurotransmitter systems, including dopamine and GABA. For example, dopamine and noradrenaline can be co-released from the same LC-noradrenergic terminals^56-58^. Locus coeruleus activity can alter midbrain dopamine cell firing^59^ and directly participate in the regulation of dopamine release in hippocampus^60^. Nevertheless, pharmacological studies using selective dopamine manipulations have found no effects on response inhibition^61-63^, and there is no clear evidence for a relationship between response inhibition and the levodopa equivalent daily dose or the on/off state of dopaminergic medication in Parkinson’s disease ^64,65^. Taken together, our results are consistent with a growing body of work that indicates a robust link between the locus-coeruleus noradrenaline system and response inhibition, although further work is needed to elucidate the potential contribution of dopaminergic mechanisms.

However, we anticipate that any future use of noradrenergic treatments that target response inhibition deficits would be adjunctive to standard dopaminergic therapy, and not an alternative. Therefore, all patients were tested on their usual clinically optimised dopaminergic medication. We acknowledge that patients’ task performance may have differed if they were taken off their dopaminergic medication, which consequently could have affected the relationship between response inhibition and locus coeruleus integrity.

Our study has several limitations. We acknowledge that all the patients were diagnosed with clinical criteria, without pathological confirmation. Misclassification of progressive supranuclear palsy subtypes and other atypical parkinsonian syndromes can occur for PSP-Richardson’s syndrome diagnosis^66^ and future studies including post-mortem confirmation would be critical to enhance the diagnostic accuracy. Data from some patients with Parkinson’s disease are drawn from a placebo-controlled drug study^26^. Only the placebo data were analysed, but we acknowledge that this might have resulted in heterogeneity due to effects of placebo expectancy and/or task practice. To mitigate these issues, we used two approaches to explicitly model the impact of placebo/practice confounds in the statistical analyses. Reassuringly, neither of these steps meaningfully altered the results.

To conclude, our study further elucidates the role of the locus coeruleus noradrenergic contribution in response inhibition and its impairment in Parkinson’s disease and Progressive Supranuclear Palsy. We propose that locus coeruleus imaging could be used as a heuristic stratification marker in clinical trials, targeting response inhibition deficits with noradrenergic drugs in those most likely to benefit. Individual differences in response to drug are marked ^26^, due to variation in disease severity, allelic variations in the noradrenaline transporter, polymorphisms in the CYP2D6 liver enzyme catabolising noradrenergic drugs, baseline brain networks structure or function, and the integrity of the locus coeruleus ^16,17,20,26,46,67,68^. Optimisation of noradrenergic treatments will benefit from better understanding the mechanisms of response inhibition and their relationship to the integrity of the locus coeruleus.

## Data Availability

The MT and MP2RAGE data in the present study are available upon reasonable request to the authors

## Acknowledgements

This study was supported by Parkinson’s UK (K-1702); the Cambridge Centre for Parkinson-Plus; the China Scholarship Council; a Neil Hamilton Fairley Fellowship from the Australian National Health and Medical Research Council (GNT1091310); a Cambridge Trust Vice-Chancellor’s Award and Fitzwilliam College Scholarship; the Association of British Neurologists – Patrick Berthoud Charitable Trust (RG99368); the Medical Research Council (SUAG/051 G101400; MR/P01271X/1); James S. McDonnell Foundation 21st Century Science Initiative Scholar Award in Understanding Human Cognition; a RCUK/UKRI Research Innovation Fellowship awarded by the Medical Research Council (MR/R007446/1); the NIHR Cambridge Clinical Research Facility and the NIHR Cambridge Biomedical Research Centre (BRC-1215-20014). The views expressed are those of the authors and not necessarily those of the NHS, the NIHR or the Department of Health and Social Care. For the purpose of open access, the author has applied a CC BY public copyright licence to any Author Accepted Manuscript version arising from this submission.

## Disclosures

The authors declared no conflict of interest.

## Notes

### Competing Interest Statement

The authors have declared no competing interest.

### Author Declarations

Cambridge Research Ethics Committees gave ethical approval for this work

