## Supplementary Materials for "Reduced locus coeruleus integrity linked to response inhibition deficits in parkinsonian disorders"

### **Supplementary** **Information**

#### LC signal validation

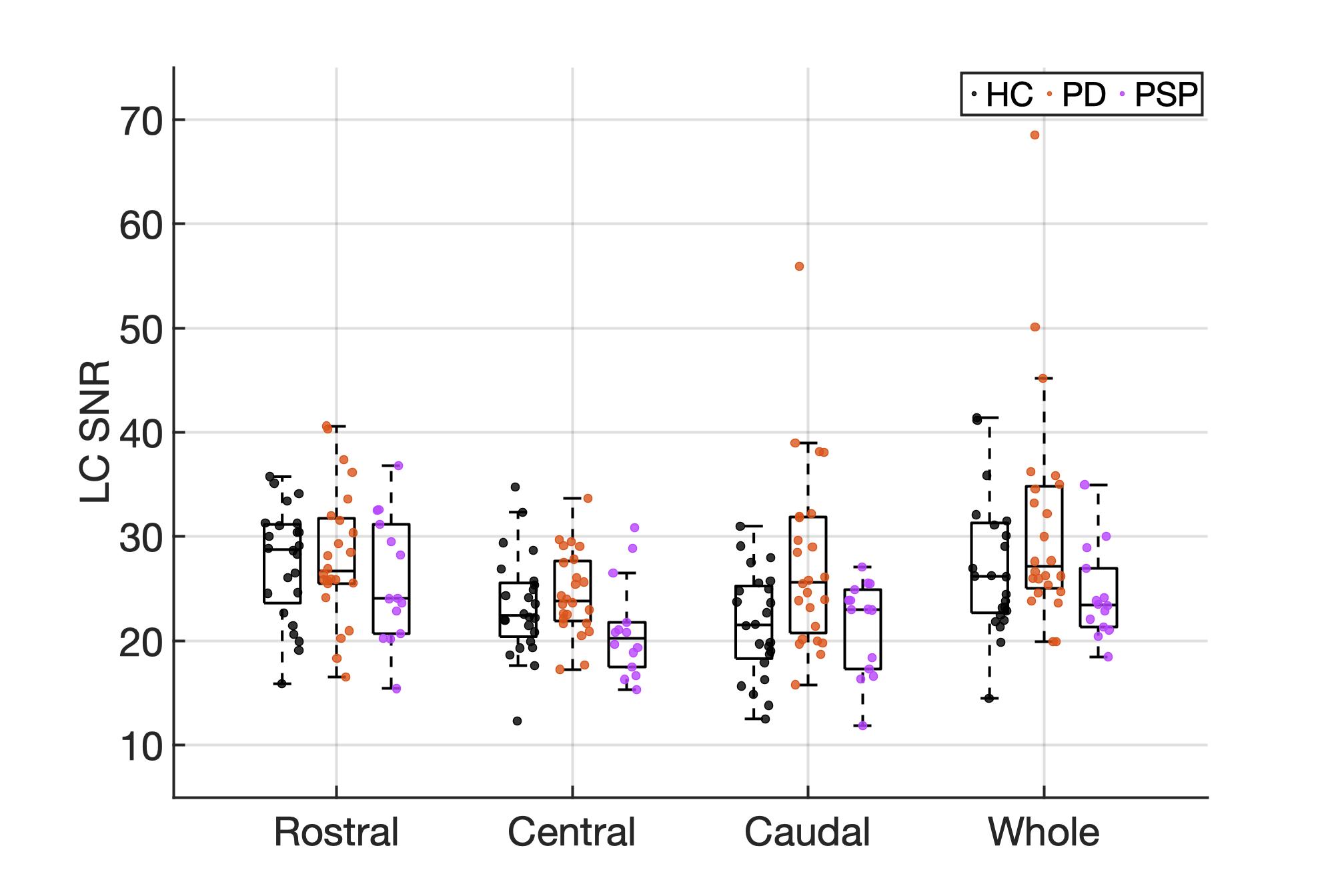

**Figure S1 | Signal-to-noise ratio (SNR) variations across LC subregions between controls, patients with PD and PSP. Repeated measures ANOVA has revealed a significant effect of LC subregions (F(2,118)=12.84, *p*<0.001), higher SNR was found in rostral LC comparing to central (*p_holm_*<0.001) and caudal (*p_holm_*=0.003) subregions. The SNR also differed across three groups (F(2,58)=4.43, *p*=0.02) where higher SNR was seen in the LC area in PD patients comparing with the other two groups (PD>PSP: *p_holm_*=0.03; PD>control: *p_holm_*=0.06). There was a trend towards a significant regions × group interaction for the SNR (F(4,116)=2.26, *p*=0.07). The group effect was not significant for the SNR measurement of the whole structure (F(2,59)=0.07, *p*=0.93).**

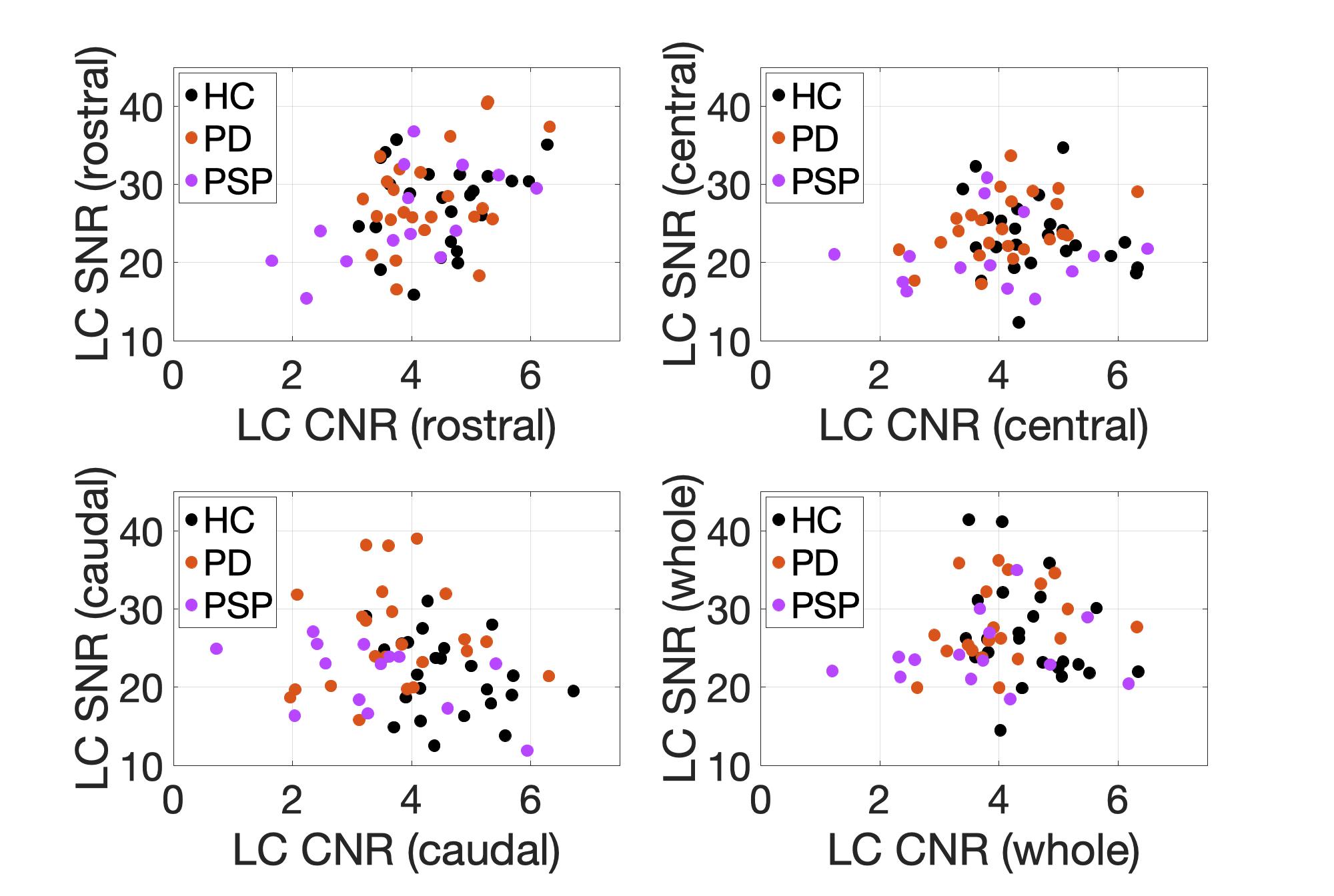

**Figure S2 | The effect of signal-to-noise ratio (SNR) on LC contrast. The atlas-extracted LC contrast in the rostral area was significantly correlated with the SNR in the same region (β=0.38, *p*=0.002). There was no correlation between the SNR and LC contrast in other subregions (central: *p*=0.59, caudal: *p*=0.1) nor for the whole LC (*p*=0.33). CNR=contrast-to-noise ratio.**

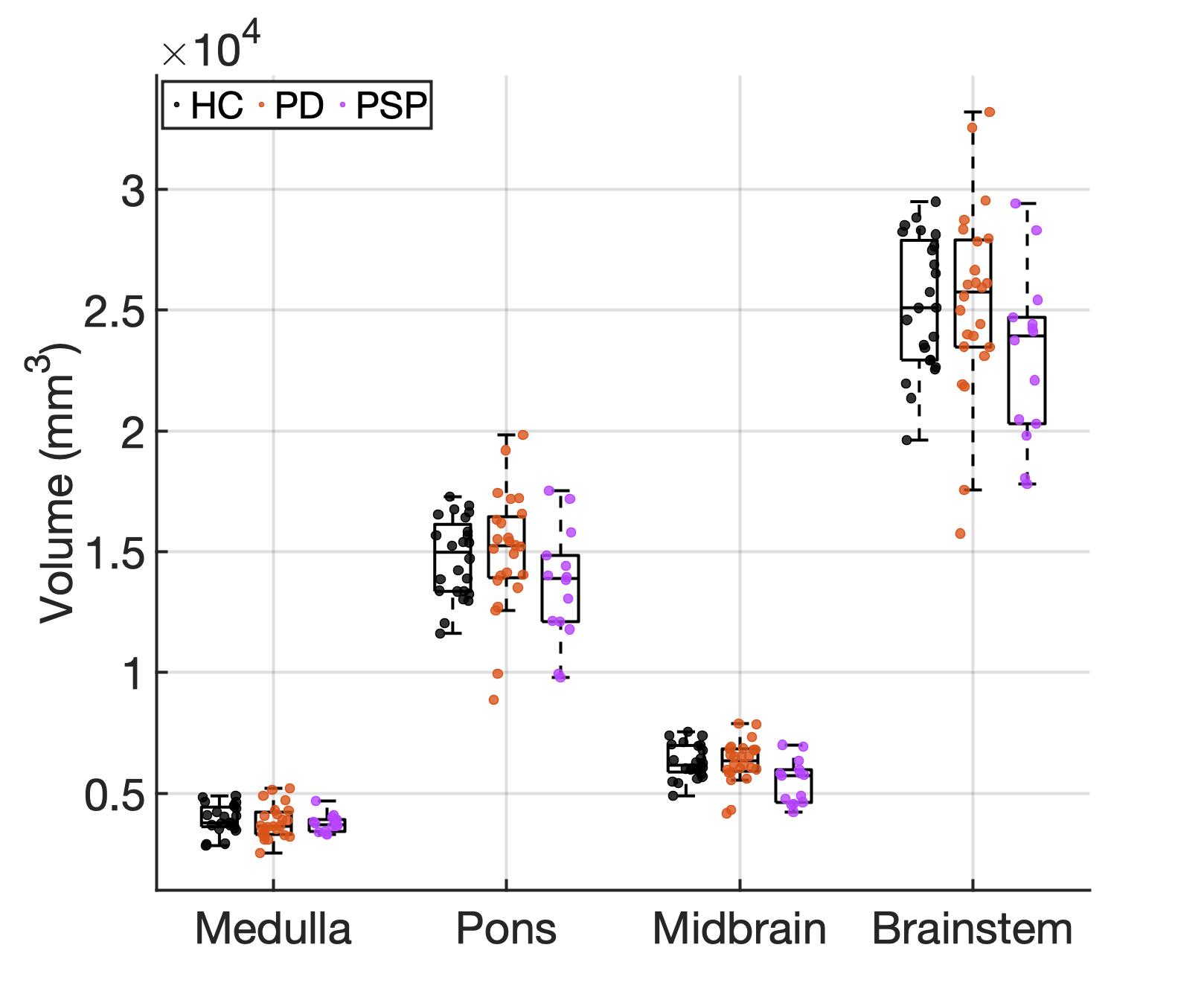

**Figure S3 | Volumetric comparisons of brainstem substructures across all groups. Disease-related brain atrophy was not found in medulla (F(2,58)=0.48, *p*=0.62), pons (F(2,58)=2, *p*=0.14) or the whole brainstem (F(2,58)=2.39, *p*=0.1). There was a significant group effect on midbrain volume (F(2,58)=5.59, *p*=0.006). Significant midbrain atrophy was found in PSP compared to control (*p_holm_*=0.008) and PD (*p_holm_*=0.01). The estimated total intracranial volume was included as covariate to control for global atrophy.**

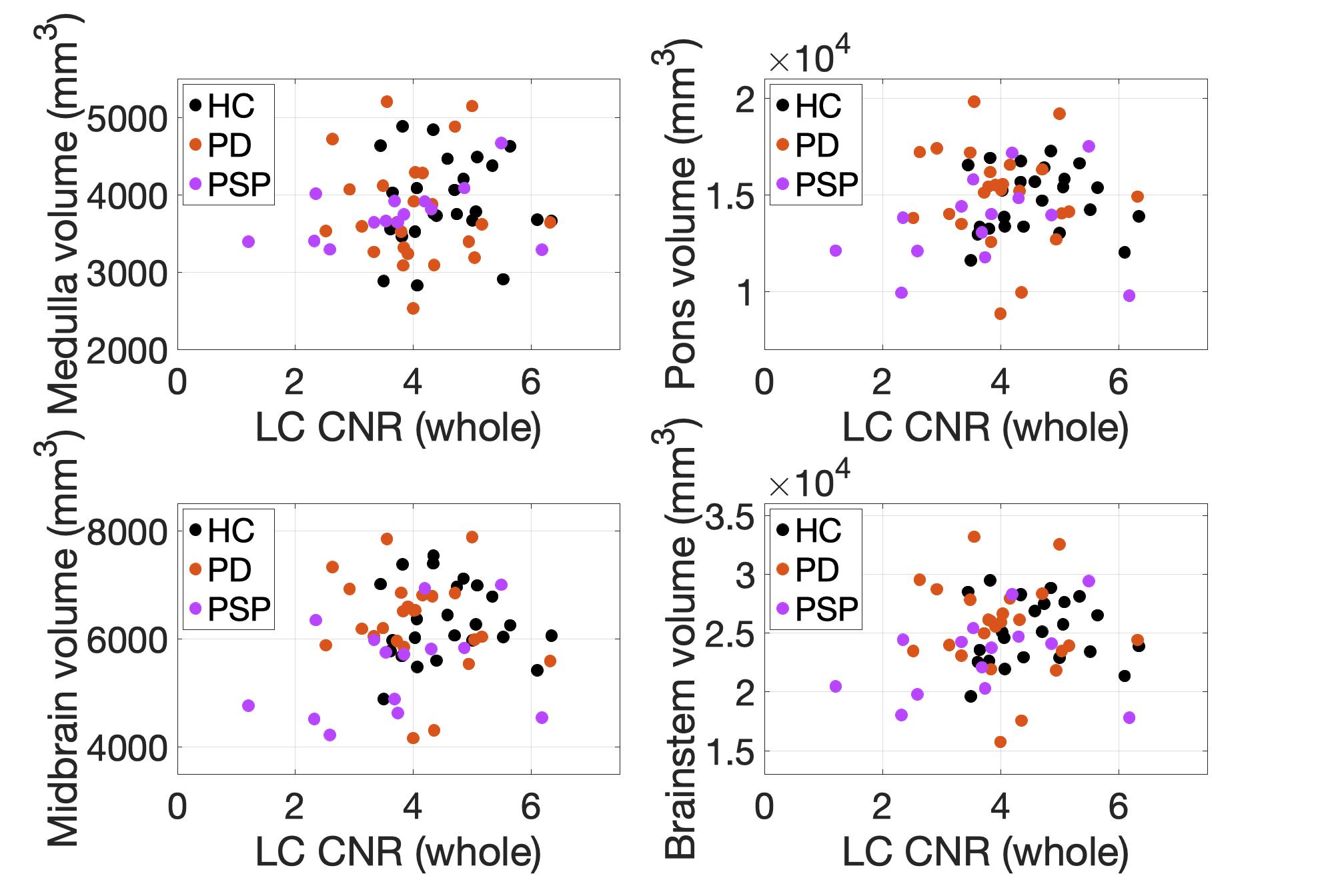

**Figure S4 | The relationship between LC contrast and brainstem atrophy. Contrast estimation was not correlated with individual brainstem volumes as examined in linear regression models with total intracranial volume as covariate (medulla: β=0.11, *p*=0.15, pons: β=0.1, *p*=0.46, midbrain: β=0.15, *p*=0.24, whole brainstem: β=0.12, *p*=0.36).**

List of priors for the race models

*Table S1: Priors for ex-Gaussian race model*

| **Parameter** | **Group-level prior distributions** | | |
| --- | --- | --- | --- |
| Mean of Gaussian component of matching go process ($\mu_{go-match}$) | $\mu_{\mu_{go-match}}$ | ~ | $\mathcal{N}_{+}\left( 1.5, 1 \right)$ |
|  | $\sigma_{\mu_{go-match}}$ | ~ | $\exp\left( 1 \right)$ |
| Mean of Gaussian component of mismatching go process ($\mu_{go-mismatch}$) | $\mu_{\mu_{go-mismatch}}$ | ~ | $\mathcal{N}_{+}\left( 1.5, 1 \right)$ |
|  | $\sigma_{\mu_{go-mismatch}}$ | ~ | $\exp\left( 1 \right)$ |
| Mean of Gaussian component of stop process ($\mu_{\mathrm{stop}}$) | $\mu_{\mu_{\mathrm{stop}}}$ | ~ | $\mathcal{N}_{+}\left( 1, 1 \right)$ |
|  | $\sigma_{\mu_{\mathrm{stop}}}$ | ~ | $\exp\left( 1 \right)$ |
| Standard deviation of Gaussian component of matching go process ($\sigma_{go-match}$) | $\mu_{\sigma_{go-match}}$ | ~ | $\mathcal{N}_{+}\left( 0.2, 1 \right)$ |
|  | $\sigma_{\sigma_{go-match}}$ | ~ | $\exp\left( 1 \right)$ |
| Standard deviation of Gaussian component of mismatching go process ($\sigma_{go-mismatch}$) | $\mu_{\sigma_{go-mismatch}}$ | ~ | $\mathcal{N}_{+}\left( 0.2, 1 \right)$ |
|  | $\sigma_{\sigma_{go-mismatch}}$ | ~ | $\exp\left( 1 \right)$ |
| Standard deviation of Gaussian component of stop process ($\sigma_{\mathrm{stop}}$) | $\mu_{\sigma_{\mathrm{stop}}}$ | ~ | $\mathcal{N}_{+}\left( 0.2, 1 \right)$ |
|  | $\sigma_{\sigma_{\mathrm{stop}}}$ | ~ | $\exp\left( 1 \right)$ |
| Mean of exponential component of matching go process ($\tau_{go-match}$) | $\mu_{\tau_{go-match}}$ | ~ | $\mathcal{N}_{+}\left( 0.2, 1 \right)$ |
|  | $\sigma_{\tau_{go-match}}$ | ~ | $\exp\left( 1 \right)$ |
| Mean of exponential component of mismatching go process ($\tau_{go-mismatch}$) | $\mu_{\tau_{go-mismatch}}$ | ~ | $\mathcal{N}_{+}\left( 0.2, 1 \right)$ |
|  | $\sigma_{\tau_{go-mismatch}}$ | ~ | $\exp\left( 1 \right)$ |
| Mean of exponential component of stop process ($\tau_{\mathrm{stop}}$) | $\mu_{\tau_{\mathrm{stop}}}$ | ~ | $\mathcal{N}_{+}\left( 0.2, 1 \right)$ |
|  | $\sigma_{\tau_{\mathrm{stop}}}$ | ~ | $\exp\left( 1 \right)$ |
| Probit transformed trigger failure probability [$\Phi^{-1}(P_{\mathrm{TF}})$] | $\mu_{\Phi^{-1}(P_{\mathrm{TF}})}$ | ~ | $\mathcal{N}\left( \Phi^{-1}(0.1), 1 \right)$ |
|  | $\sigma_{\Phi^{-1}(P_{\mathrm{TF}})}$ | ~ | $\exp\left( 1 \right)$ |
| Probit transformed go failure probability [$\Phi^{-1}(P_{\mathrm{GF}})$] | $\mu_{\Phi^{-1}(P_{\mathrm{GF}})}$ | ~ | $\mathcal{N}\left( \Phi^{-1}\left( 0.1 \right), 1 \right)$ |
|  | $\sigma_{\Phi^{-1}(P_{\mathrm{GF}})}$ | ~ | $\exp\left( 1 \right)$ |

Where $\mathcal{N(}\mu,\sigma)$ denotes a normal distribution with mean $\mu$ and standard deviation $\sigma$; $\mathcal{N}_{+}(\mu,\sigma)$ denotes a normal distribution truncated to only allow positive values; $exp(\lambda)$ denotes an exponential distribution with rate parameter $\lambda$; and $\Phi^{-1}(p)$ denotes the probit function (i.e., the inverse cumulative distribution function of the standard normal distribution) evaluated at probability $p$. All parameters are on the scale of seconds, except for the trigger and go failure probabilities.

*Table S2: Priors for hybrid Wald / ex-Gaussian race model*

| **Parameter** | **Group-level prior distributions** | | |
| --- | --- | --- | --- |
| Drift rate of matching go process ($v_{go-match}$) | $\mu_{v_{go-match}}$ | ~ | $\mathcal{N}_{+}\left( 2, 3 \right)$ |
|  | $\sigma_{v_{go-match}}$ | ~ | $\exp\left( 1 \right)$ |
| Drift rate of mismatching go process ($v_{go-mismatch}$) | $\mu_{v_{go-mismatch}}$ | ~ | $\mathcal{N}_{+}\left( 1, 3 \right)$ |
|  | $\sigma_{v_{go-mismatch}}$ | ~ | $\exp\left( 1 \right)$ |
| Threshold of go processes ($B_{\mathrm{go}}$) | $\mu_{B_{\mathrm{go}}}$ | ~ | $\mathcal{N}_{+}\left( 2, 1 \right)$ |
|  | $\sigma_{B_{\mathrm{go}}}$ | ~ | $\exp\left( 1 \right)$ |
| Non-decision time of go processes (${t_{0}}_{\mathrm{go}}$) | $\mu_{{t_{0}}_{\mathrm{go}}}$ | ~ | $\mathcal{N}_{(0.1,\infty)}\left( 0.3, 0.25 \right)$ |
|  | $\sigma_{{t_{0}}_{\mathrm{go}}}$ | ~ | $\exp\left( 1 \right)$ |
| Mean of Gaussian component of stop process ($\mu_{\mathrm{stop}}$) | $\mu_{\mu_{\mathrm{stop}}}$ | ~ | $\mathcal{N}_{+}\left( 1, 1 \right)$ |
|  | $\sigma_{\mu_{\mathrm{stop}}}$ | ~ | $\exp\left( 1 \right)$ |
| Standard deviation of Gaussian component of stop process ($\sigma_{\mathrm{stop}}$) | $\mu_{\sigma_{\mathrm{stop}}}$ | ~ | $\mathcal{N}_{+}\left( 0.2, 1 \right)$ |
|  | $\sigma_{\sigma_{\mathrm{stop}}}$ | ~ | $\exp\left( 1 \right)$ |
| Mean of exponential component of stop process ($\tau_{\mathrm{stop}}$) | $\mu_{\tau_{\mathrm{stop}}}$ | ~ | $\mathcal{N}_{+}\left( 0.2, 1 \right)$ |
|  | $\sigma_{\tau_{\mathrm{stop}}}$ | ~ | $\exp\left( 1 \right)$ |
| Probit transformed trigger failure probability [$\Phi^{-1}(P_{\mathrm{TF}})$] | $\mu_{\Phi^{-1}(P_{\mathrm{TF}})}$ | ~ | $\mathcal{N}\left( \Phi^{-1}(0.1), 1 \right)$ |
|  | $\sigma_{\Phi^{-1}(P_{\mathrm{TF}})}$ | ~ | $\exp\left( 1 \right)$ |
| Probit transformed go failure probability [$\Phi^{-1}(P_{\mathrm{GF}})$] | $\mu_{\Phi^{-1}(P_{\mathrm{GF}})}$ | ~ | $\mathcal{N}\left( \Phi^{-1}\left( 0.1 \right), 1 \right)$ |
|  | $\sigma_{\Phi^{-1}(P_{\mathrm{GF}})}$ | ~ | $\exp\left( 1 \right)$ |

Where $\mathcal{N(}\mu,\sigma)$ denotes a normal distribution with mean $\mu$ and standard deviation $\sigma$; $\mathcal{N}_{+}(\mu,\sigma)$ denotes a normal distribution truncated to only allow positive values; $\mathcal{N}_{(a,b)}(\mu,\sigma)$ denotes a normal distribution with lower truncation $a$ and upper truncation $b$; $exp(\lambda)$ denotes an exponential distribution with rate parameter $\lambda$; and $\Phi^{-1}(p)$ denotes the probit function (i.e., the inverse cumulative distribution function of the standard normal distribution) evaluated at probability $p$. The drift rates and threshold are on an arbitrary scale; the non-decision time and the ex-Gaussian parameters are on the scale of seconds.

#### Goodness of fit for the ex-Gaussian model

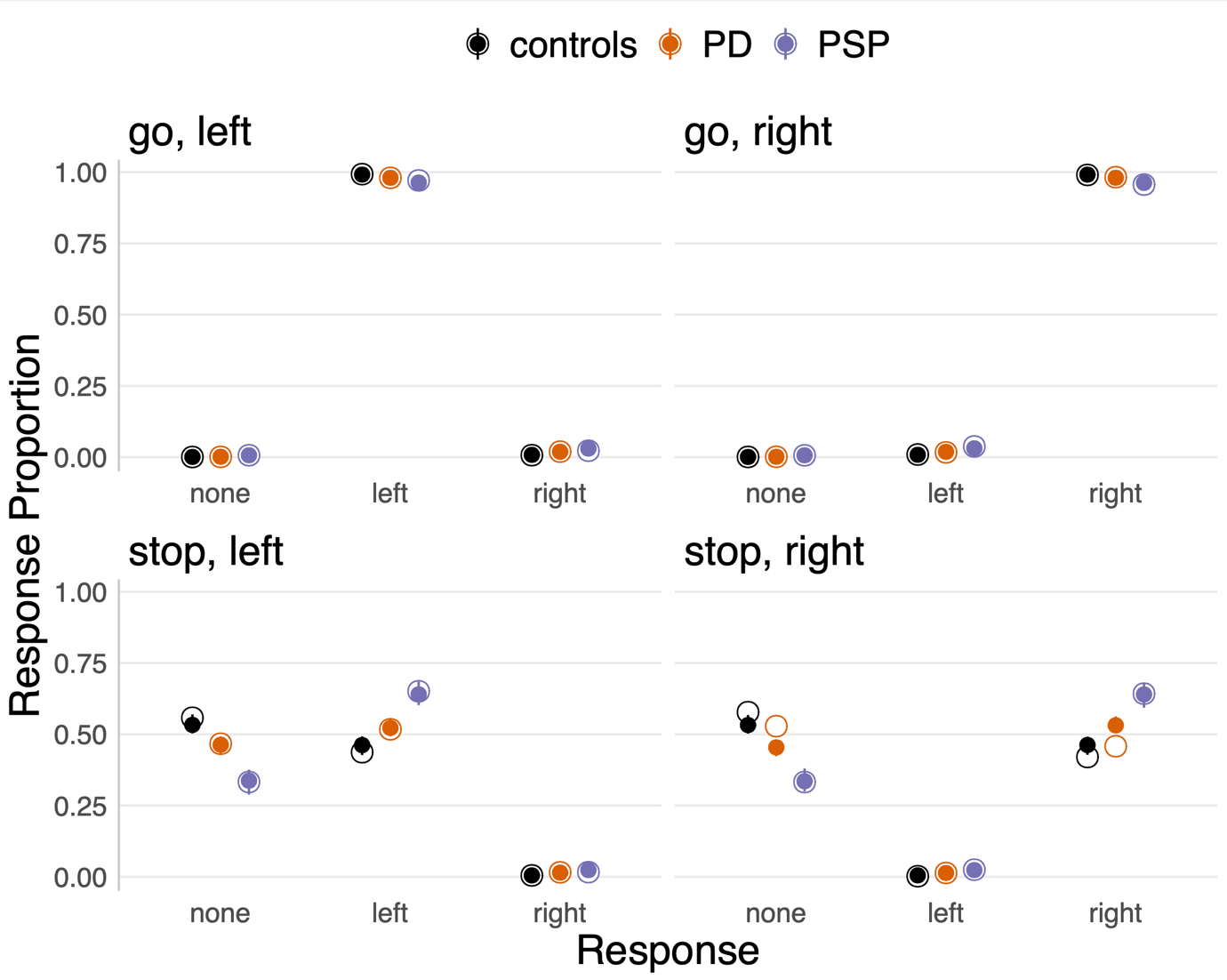

**Figure S5 | Posterior predictive check of response proportions. The observed data (hollow dots) is compared with simulated data from the final model fit (solid dots). Each panel represents one unique combination of trial type (go versus stop) and choice stimulus (left versus right). Within each panel, the observed group-level median response proportions are plotted separately for each response option (no response, left, right) and group (controls, PD, PSP). The model simulations are illustrated as the median (solid dot) and 95% quantile interval (error bar) of 100 simulated participants, randomly drawn from the posterior predictive distribution.**

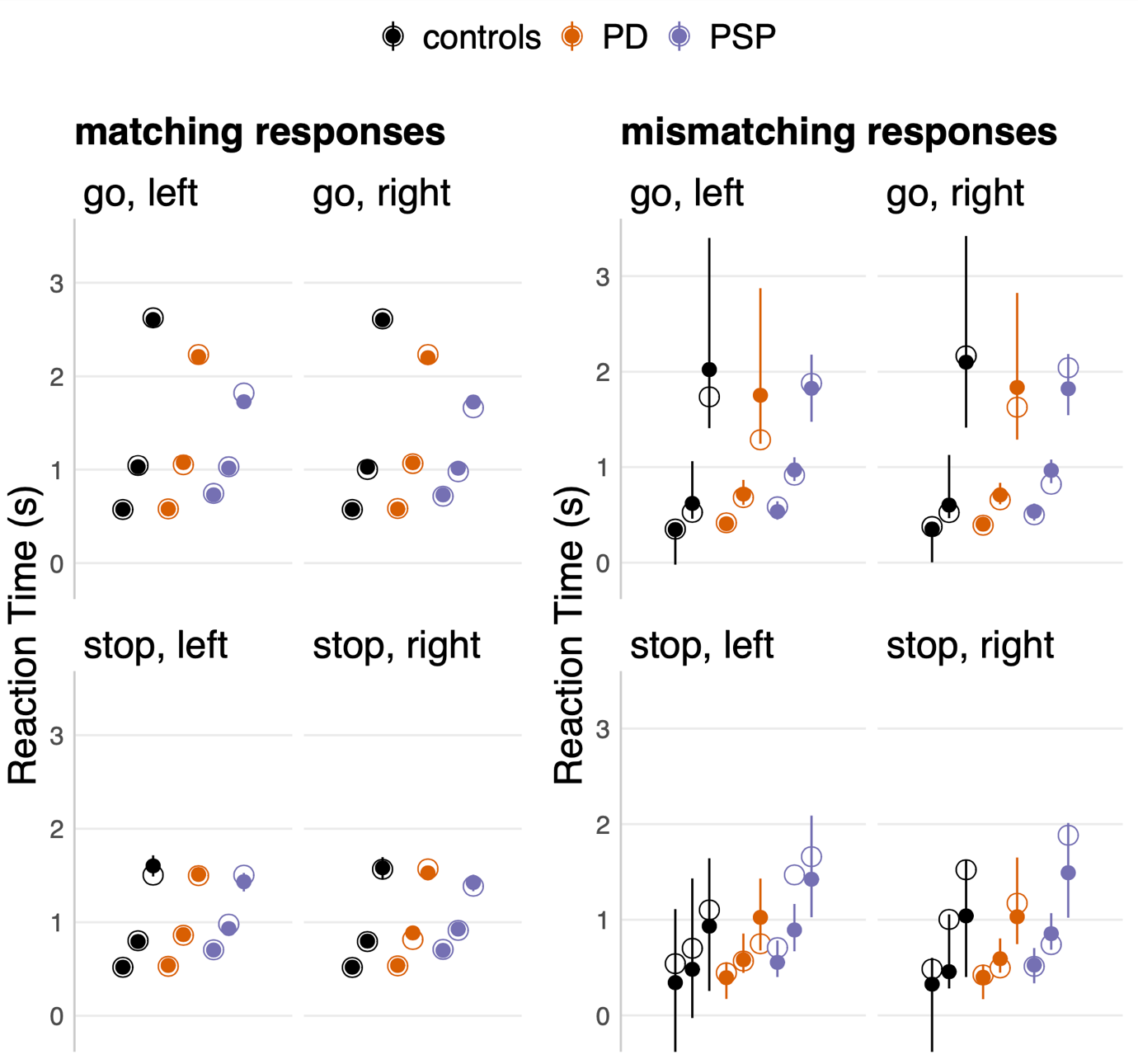

**Figure S6 | Posterior predictive check of reaction times. The observed data (hollow dots) is compared with simulated data from the final model fit (solid dots). Each panel represents one unique combination of trial type (go versus stop) and choice stimulus (left versus right). Within each panel, the lower, middle and upper set of dots represent the 10^th^, 50^th^, and 90^th^ percentiles of the reaction time distributions, respectively. The model simulations are illustrated as the median (solid dot) and 95% quantile interval (error bar) of 100 simulated participants, randomly drawn from the posterior predictive distribution. The posterior predictive check is illustrated separately for matching (correct) responses and mismatching (incorrect) responses, as mismatching responses constituted a very small proportion of the data (Figure 2D).**

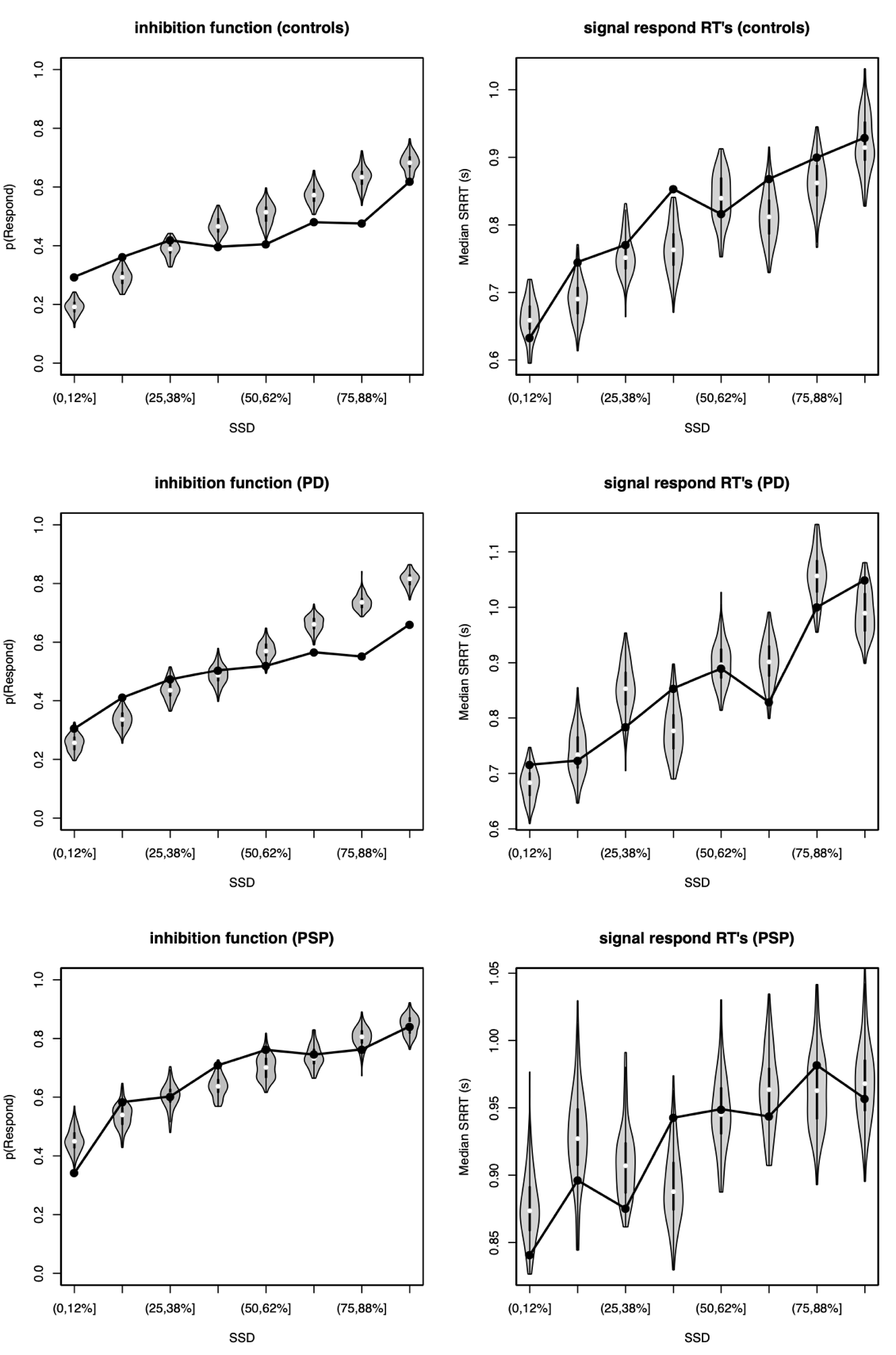

**Figure S7 | Posterior predictive check of stopping performance. The observed data (black dots and lines) is compared with simulated data from the final model fit (grey violin plots). The panels in the left column illustrate the mean probability of responding (i.e., mean probability of stop failure) as a function of the stop signal delay (SSD). The panels in the right column illustrate the median reaction time for stop trials (i.e., median signal respond RT), as a function of the SSD. The SSD was binned by percentile ranges. The model simulations are illustrated as the median (white solid dot) and full distribution (grey violin plot) of 100 simulated participants, randomly drawn from the posterior predictive distribution.**

#### Selection of predictors of response inhibition

*Table S3: Stepwise selection of predictors of response inhibition scores for the full sample (controls, PD and PSP groups).*

|  | **Model selection step** | | | | | | | | | | | | | | |
| --- | --- | --- | --- | --- | --- | --- | --- | --- | --- | --- | --- | --- | --- | --- | --- |
| **Predictors** | 1 | | 2 | | 3 | | 4 | | | 5 | | 6 | | 7 | 8 |
| LV_LC_ | 0.25^*^ | | 0.24^*^ | | 0.24^*^ | | 0.24^*^ | | | 0.23^*^ | | 0.24^**^ | | 0.24^**^ | 0.24^**^ |
| Group | -0.71^***^; -0.37^**^ | | -0.69^***^; -0.36^**^ | | -0.68^***^; -0.36^**^ | | -0.67^***^; -0.37^**^ | | | -0.68^***^; -0.36^**^ | | -0.70^***^; -0.36^**^ | | -0.69^***^; -0.35^**^ | -0.68^***^; -0.37^**^ |
| Gender | 0.12 | | 0.14 | | 0.14 | | 0.14 | | | 0.11 | | 0.10 | | 0.07 | - |
| Brainstem | 0.40 | | 0.37 | | 0.37 | | 0.10 | | | 0.09 | | 0.07 | | - | - |
| Education | -0.08 | | -0.08 | | -0.07 | | -0.08 | | | -0.07 | | - | | - | - |
| TIV | 0.06 | | 0.07 | | 0.06 | | 0.07 | | | - | | - | | - | - |
| Pons | -0.31 | | -0.28 | | -0.28 | | - | | | - | | - | | - | - |
| Age | -0.04 | | -0.03 | | - | | - | | | - | | - | | - | - |
| LV_LC_ × Group | -0.04; 0.08 | | - - | | - - | | - - | | | - - | | - - | | - - | - - |
| **Information Criteria** | |  | |  | |  | |  |  | |  | |  | | |
| AIC | 138.11 | | 134.52 | | 132.69 | | 131.03 | | | 129.71 | | 128.43 | | 126.98 | 125.78 |
| ∆ AIC | 12.33 | | 8.74 | | 6.91 | | 5.25 | | | 3.94 | | 2.65 | | 1.20 | 0 |
| BIC | 165.76 | | 157.92 | | 153.96 | | 150.18 | | | 146.73 | | 143.32 | | 139.74 | 136.41 |
| ∆ BIC | 29.35 | | 21.51 | | 17.55 | | 13.76 | | | 10.32 | | 6.90 | | 3.32 | 0 |

*Note*. Values for predictors are standardised regression coefficients (ß). ^***^*p* < .001; ^**^*p* < .01; ^*^ *p* < .05. LV_LC_: Participant scores on latent variable of locus coeruleus CNR; Group: controls vs. PD patients vs. PSP patients; Brainstem: Total brainstem volume (mm^3^); Education: Years of education; TIV: Estimated total intracranial volume (mm^3^); Pons: Pons volume (mm^3^); AIC: Akaike Information Criterion; BIC: Bayesian Information Criterion; ∆ AIC / BIC: difference in AIC / BIC with respect to the lowest AIC / BIC value. Note that the predictors Group and LV_LC_ × Group are each represented by two regression coefficients: The first coefficient represents the difference between the control group and the intercept; the second coefficient represents the difference between the PD group and the intercept; and the PSP group is modelled as the intercept plus the negative of the sum of the two coefficients.

*Table S4: Inclusion probabilities and Bayes Factors for the inclusion of predictors of response inhibition scores for the full sample (controls, PD and PSP groups).*

| Predictors | P(incl) | P(incl\|data) | P(excl\|data) | BF_inclusion_ |
| --- | --- | --- | --- | --- |
| LV_LC_ | 0.40 | 0.73 | 0.09 | 7.44 |
| Group | 0.40 | 0.83 | 7.71 × 10^-8^ | 1.08 × 10^7^ |
| Gender | 0.50 | 0.29 | 0.71 | 0.41 |
| Brainstem | 0.50 | 0.31 | 0.69 | 0.45 |
| Education | 0.50 | 0.33 | 0.67 | 0.48 |
| TIV | 0.50 | 0.30 | 0.70 | 0.43 |
| Pons | 0.50 | 0.30 | 0.70 | 0.43 |
| Age | 0.50 | 0.29 | 0.71 | 0.40 |
| LV_LC_ × Group | 0.20 | 0.17 | 0.73 | 0.23 |

Note. P(incl): prior inclusion probability, i.e. the summed prior probability of models that include the predictor. A priori, all possible restrictions of the full model were deemed to be equally likely (i.e., a uniform prior was assigned to the model space). Thus, P(incl) reflects the proportion of alternative models that included the predictor. P(incl|data): posterior inclusion probability, i.e. the summed posterior probability of models that include the predictor. P(excl|data): posterior exclusion probability, i.e. the summed posterior probability of models that exclude the predictor. BF_inclusion_: Inclusion Bayes Factor, i.e. the change from prior to posterior inclusion odds. This indicates how much more likely the data are under models that include the predictor, compared to models that exclude the predictor (1). This analysis was performed using “matched” models, which means that (i) models were not permitted to include an interaction effect without its constituent main effects, and (ii) inclusion probabilities for an interaction effect were based only on the subset of models that contained (at least) the constituent main effects of the interaction.

*Table S5: Stepwise selection of predictors of response inhibition scores for the patients only (PD and PSP groups).*

|  | **Model selection step** | | | | | | | | | | | | | | | | | | |
| --- | --- | --- | --- | --- | --- | --- | --- | --- | --- | --- | --- | --- | --- | --- | --- | --- | --- | --- | --- |
| **Predictors** | 1 | | 2 | | 3 | | 4 | | 5 | | 6 | | 7 | | 8 | | 9 | | 10 |
| LV_LC_ | 0.24 | | 0.23 | | 0.22 | | 0.23 | | 0.23 | | 0.24 | | 0.24^*^ | | 0.25^*^ | | 0.25^*^ | | 0.26^*^ |
| Group | -0.60^***^ | | -0.60^***^ | | -0.59^***^ | | -0.59^***^ | | -0.60^***^ | | -0.60^***^ | | -0.58^***^ | | -0.59^***^ | | -0.61^***^ | | -0.66^***^ |
| Education | -0.16 | | -0.16 | | -0.16 | | -0.16 | | -0.17 | | -0.17 | | -0.15 | | -0.16 | | -0.14 | | - |
| Dx years | -0.11 | | -0.12 | | -0.12 | | -0.13 | | -0.12 | | -0.11 | | -0.11 | | -0.11 | | - | | - |
| Gender | 0.20 | | 0.20 | | 0.22 | | 0.22 | | 0.22 | | 0.15 | | 0.10 | | - | | - | | - |
| Brainstem | 0.38 | | 0.16 | | 0.16 | | 0.16 | | 0.14 | | 0.11 | | - | | - | | - | | - |
| TIV | 0.08 | | 0.08 | | 0.09 | | 0.09 | | 0.10 | | - | | - | | - | | - | | - |
| UPDRS III | 0.11 | | 0.11 | | 0.09 | | 0.08 | | - | | - | | - | | - | | - | | - |
| Age | -0.06 | | -0.05 | | -0.04 | | - | | - | | - | | - | | - | | - | | - |
| LV_LC_ × Group | 0.05 | | 0.04 | | - | | - | | - | | - | | - | | - | | - | | - |
| Pons | -0.22 | | - | | - | | - | | - | | - | | - | | - | | - | | - |
| **Information Criteria** | |  | |  | |  | |  | |  | |  | |  | |  | |  | |
| AIC | 98.08 | | 96.16 | | 94.27 | | 92.35 | | 90.79 | | 89.47 | | 88.20 | | 87.01 | | 85.99 | | 85.55 |
| ∆ AIC | 12.54 | | 10.61 | | 8.72 | | 6.81 | | 5.24 | | 3.92 | | 2.65 | | 1.46 | | 0.44 | | 0 |
| BIC | 119.37 | | 115.81 | | 112.28 | | 108.73 | | 105.53 | | 102.57 | | 99.66 | | 96.83 | | 94.18 | | 92.10 |
| ∆ BIC | 27.27 | | 23.71 | | 20.18 | | 16.63 | | 13.43 | | 10.47 | | 7.56 | | 4.73 | | 2.08 | | 0 |

*Note*. Values for predictors are standardised regression coefficients (ß). ^***^*p* < .001; ^**^*p* < .01; ^*^ *p* < .05. LV_LC_: Participant scores on latent variable of locus coeruleus CNR; Group: PD patients vs. PSP patients; Education: Years of education; Dx years: Years since diagnosis; Brainstem: total brainstem volume (mm^3^); TIV: Estimated total intracranial volume (mm^3^); UPDRS III: Unified Parkinson’s Disease Rating Scale, motor examination; Pons: pons volume; AIC: Akaike Information Criterion; BIC: Bayesian Information Criterion; ∆ AIC / BIC: difference in AIC / BIC with respect to the lowest AIC / BIC value.

*Table S6: Inclusion probabilities and Bayes Factors for the inclusion of predictors of response inhibition scores for the patients only (PD and PSP groups).*

| Predictors | P(incl) | P(incl\|data) | P(excl\|data) | BF_inclusion_ |
| --- | --- | --- | --- | --- |
| LV_LC_ | 0.40 | 0.50 | 0.22 | 2.31 |
| Group | 0.40 | 0.72 | 3.39 × 10^-4^ | 2108.87 |
| Education | 0.50 | 0.47 | 0.53 | 0.89 |
| Dx years | 0.50 | 0.41 | 0.59 | 0.68 |
| Gender | 0.50 | 0.36 | 0.64 | 0.56 |
| Brainstem | 0.50 | 0.36 | 0.64 | 0.55 |
| TIV | 0.50 | 0.35 | 0.65 | 0.54 |
| UPDRS III | 0.50 | 0.38 | 0.62 | 0.61 |
| Age | 0.50 | 0.34 | 0.66 | 0.52 |
| LV_LC_ × Group | 0.20 | 0.28 | 0.50 | 0.57 |
| Pons | 0.50 | 0.35 | 0.65 | 0.55 |

Note. P(incl): prior inclusion probability, i.e. the summed prior probability of models that include the predictor. A priori, all possible restrictions of the full model were deemed to be equally likely (i.e., a uniform prior was assigned to the model space). Thus, P(incl) reflects the proportion of alternative models that included the predictor. P(incl|data): posterior inclusion probability, i.e. the summed posterior probability of models that include the predictor. P(excl|data): posterior exclusion probability, i.e. the summed posterior probability of models that exclude the predictor. BF_inclusion_: Inclusion Bayes Factor, i.e. the change from prior to posterior inclusion odds. This indicates how much more likely the data are under models that include the predictor, compared to models that exclude the predictor (1). This analysis was performed using “matched” models, which means that (i) models were not permitted to include an interaction effect without its constituent main effects, and (ii) inclusion probabilities for an interaction effect were based only on the subset of models that contained (at least) the constituent main effects of the interaction.

*Table S7: Results of regression model including practice as a covariate*

| Predictors | ß | *F* | *p* | BF_inclusion_ |
| --- | --- | --- | --- | --- |
| LV_LC_ | 0.24 | 7.11 | .010 | 6.86 |
| Group | -0.69; -0.39 | 29.12 | < .001 | 1.95 × 10^7^ |
| LV_LC_ × Group | -0.07; 0.10 | 0.31 | .822 | 0.15 |
| Practice | -0.03 | 0.05 | .737 | 0.39 |

*Note*. LV_LC_: Participant scores on latent variable of locus coeruleus CNR; Group: controls vs. PD patients vs. PSP patients; Practice: single / first session vs. second session; BF_inclusion_: inclusion Bayes Factor, i.e. the change from prior to posterior inclusion odds. For Group and LV_LC_ × Group, the first coefficient represents the difference between the control group and the intercept; the second coefficient represents the difference between the PD group and the intercept; and the PSP group is modelled as the intercept plus the negative of the sum of the two coefficients.

*Table S8: Results of regression model including placebo administration and practice as a covariate*

| Predictors | ß | *F* | *p* | BF_inclusion_ |
| --- | --- | --- | --- | --- |
| LV_LC_ | 0.24 | 6.65 | .013 | 6.75 |
| Group | -0.73; -0.31 | 27.36 | < .001 | 1.88 × 10^7^ |
| LV_LC_ × Group | -0.06; 0.09 | 0.24 | .787 | 0.15 |
| Practice & Placebo | 0.08; -0.10 | 0.17 | .847 | 0.29 |

*Note*. LV_LC_: Participant scores on latent variable of locus coeruleus CNR; Group: controls vs. PD patients vs. PSP patients; Practice & Placebo: single session without placebo vs. first session on placebo vs. second session on placebo; BF_inclusion_: inclusion Bayes Factor, i.e. the change from prior to posterior inclusion odds. For Group and LV_LC_ × Group, the first coefficient represents the difference between the control group and the intercept; the second coefficient represents the difference between the PD group and the intercept; and the PSP group is modelled as the intercept plus the negative of the sum of the two coefficients. For Practice & Placebo, the first coefficient represents the difference between the group of participants who performed a single session without placebo and the intercept; the second coefficient represents the difference between the group of participants who were on placebo for their first session and the intercept; and the group of participants who were on placebo for their second session is modelled as the intercept plus the negative of the sum of the two coefficients.
